# Systematic and Bibliometric Insights of Over Seven Decades of Sickle Cell Disease Research in Africa

**DOI:** 10.1101/2025.10.14.25338027

**Authors:** Emohchonne Utos Jonathan, Dauda Wadzani Palnam, Peter Abraham, Israel Ogwuche Ogra, Seun Cecilia Joshua, Morumda Daji, Dasoem Naanswan Joseph, Dogara Elisha Tumba, Mela Ilu Luka, Okechukwu Kalu Iroha, Grace Peter Wabba, Ndukwe K. Johnson, Mercy Nathaniel, Elkanah Glen, Samson Usman, Emmanuel Oluwadare Balogun, Umezuruike Linus Opara

## Abstract

Sickle cell disease (SCD) remains a major public health challenge in Africa, yet the research landscape is often fragmented, with limited policy translation. This study offers a comprehensive review of SCD research in Africa over seventy-four (74) years, integrating a systematic literature review and bibliometric analysis to assess trends, influence, and collaboration patterns. A total of 2,132 articles published between 1926 and 2024 were retrieved from the Scopus database using PRISMA guidelines. Bibliometric tools including VOSviewer and the R-based Bibliometrix package enabled citation analysis, co-authorship mapping, and keyword clustering. The findings reveal 7,178 contributing authors across 728 journals, with an average of 11.16 citations per publication. The year 2022 recorded the highest output, with 167 articles, while 1970 had the highest average number of citations per article. Williams, TN was the most cited author, while Makani J. emerged as the most prolific and impactful. Muhimbili University of Health and Allied Sciences in Tanzania led in institutional productivity, and Nigeria–Niger collaborations were the most frequent. Research trends have shifted from early clinical understanding toward management innovations and therapeutic development. However, critical gaps persist in multi-omics integration, access to newborn screening, regional trial participation, and implementation science. The study calls for an inclusive research strategy that aligns with national health policies, boosts regional collaboration, and ensures evidence-based interventions for SCD across Africa.

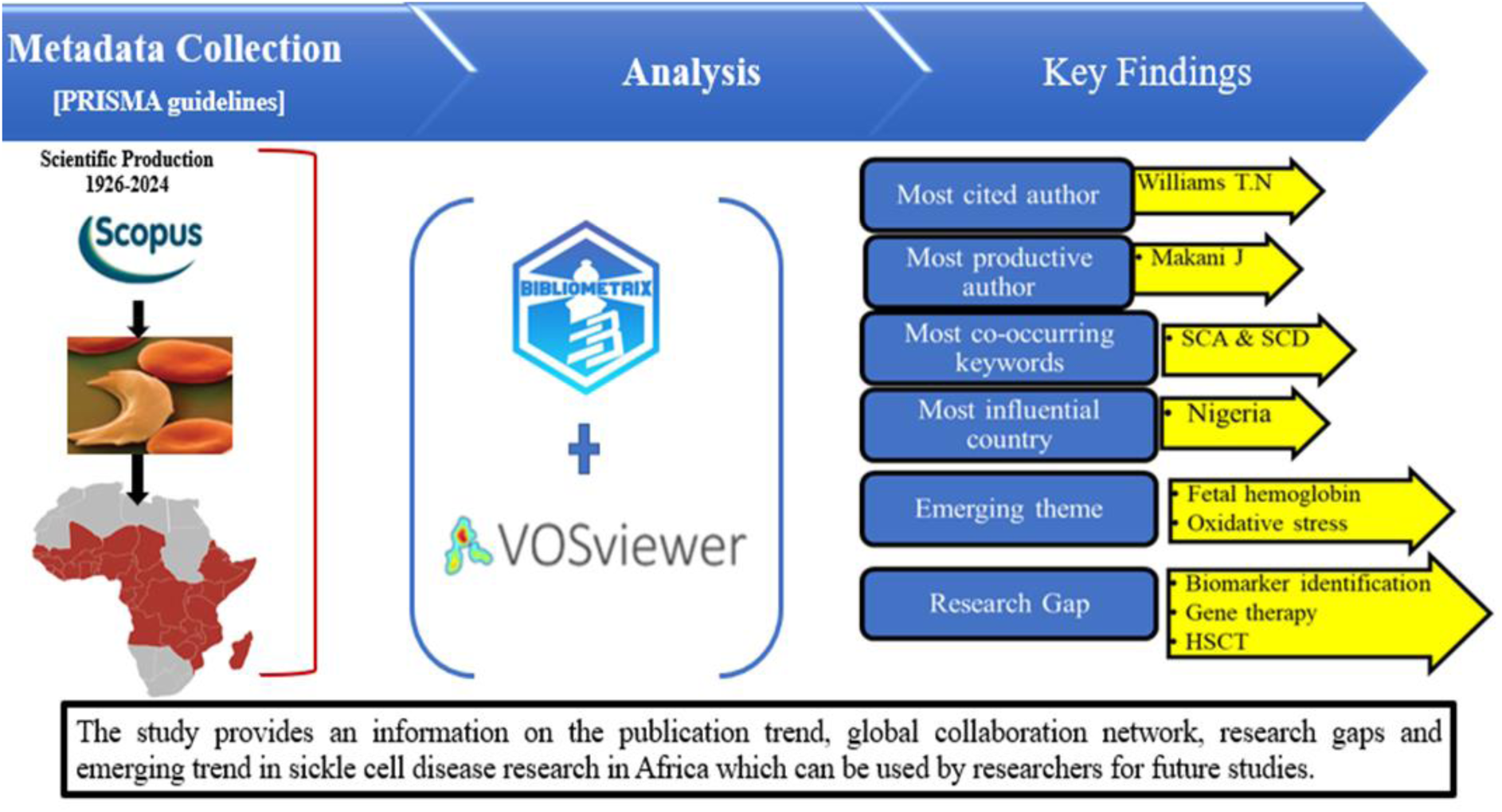

## 1.0 INTRODUCTION

Sickle cell disease (SCD) is a group of genetic disorders of blood that affect the structure of hemoglobin molecules which causes erythrocytes to assume a sickle shape. It is characterized by abnormal hemoglobin, known as hemoglobin S (HbS) (Hulihan *et al*., 2017). Sickle cell disease represents a chain of causes and effects; a mutation of a single nucleotide in the β-globin gene causes the substitution of one amino acid in the β-globin protein. This amino acid replacement causes sickle cell hemoglobin (HbS) molecules to polymerize. Consequently, hemoglobin polymers alter the membrane function and shape of the cells. These changes eventually lead to obstruction of blood vessels and increased hemolysis, which are the causes of the symptoms and complications of the disease (Conran & de Paula, 2020; Nader *et al*., 2020). Sickled RBCs become rigid, stiff, and fragile (Nader *et al*., 2020), thus altering the normal flexible biconcave shape (Damanhouri *et al.,* 2015), and are therefore highly susceptible to hemolysis. SCD is the most common inherited disease in the world and one of the top fifty (50) most common causes of death from non-communicable diseases (Piel *et al*., 2023). SCD predominantly affects individuals of African, Middle Eastern, Indian, and Mediterranean ancestry (Oron *et al.,* 2020) and is highly prevalent in regions where the sickle trait is common (Okoroiwu *et al.,* 2020).

The greatest burden of the disease is in Africa with approximately 80% of global cases (Inusa *et al.,* 2019). The prevalence of SCD in Africa represents a significant public health concern as it impacts millions of people across the continent. The unique environmental and genetic factors that are prevalent in various African regions exacerbate the disease burden thereby making it critical to understand the social, economic and health related implications of the disease. The disease poses significant challenges to affected individuals and their caregivers. The burden is heavy in countries such as Nigeria, the Democratic Republic of Congo, Uganda, and Tanzania (Akinsete, 2022; Piel *et al.,* 2023) which are home to the largest proportion of individuals afflicted by the disease (Inusa *et al.,* 2019).

Despite its public health significance, research on SCD in Africa remains disproportionately low compared to its impact (Musa *et al*., 2021). Recently, there has been a growing need to strengthen research efforts to address the unique challenges of SCD in Africa which include improving neonatal screening, understanding the genetic variation, management/treatment access, and addressing socioeconomic factors that exacerbate the disease burden. In view of rising awareness and research efforts, a bibliometric analysis of scholarly literature on SCD in Africa will reveal trends in publications, authorship and collaboration networks.

Bibliometric analysis is a statistical study of scholarly articles that uses both qualitative and quantitative methods to analyze the content, references, citations and co-authorship of publications (Fu *et al*., 2023, Rojas-Sánchez *et al.,* 2023). It is a systematic study that is carried out to identify patterns, trends and impact within a certain field. This technique is increasingly becoming popular for assessing vast number of scientific literatures (Passas, 2024) and has become an important tool for measuring scientific outputs of any research field as it examines how the intellectual, social, and conceptual structure of a particular field has evolved overtime based on the relationship and interactions between various scientific items [papers, authors, keywords, source, institution, and countries] (Öztürk *et al.,* 2024). The emergence, availability and accessibility of bibliometric tools such as VOSviewer, CiteSpace, R package ‘bibliometrix’, BibExcel, Histcite and scientific database such as Scopus, web of science, PubMed, Dimensions, Lens and Google scholar are responsible for the benefits and conveniences that bibliometric analysis offers. In the academic sphere, bibliometric analysis has become an instrumental tool that is used for conducting systematic literature review (Lim & Kumar, 2024). Bibliometric analysis provides an opportunity to systematically explore the state of SCD research in Africa. It can also provide insights into the productivity, impact and structure of research on SCD in Africa. By examining publication trends, collaboration networks, trending topics, and thematic evolution, policymakers and researchers can identify gaps and prioritize research in those areas.

While global and country-specific bibliometric analysis on SCD exist, a search of the literature revealed that few focus explicitly on Africa. Adigwe & Onoja, (2023) provided a critical review of sickle cell disease burden and challenges in Sub-Saharan Africa from 2003 to 2021 and Adesina & Opesade, (2018) conducted a bibliometric analysis of sickle cell anaemia literature on Nigeria listed in PubMed between 2006 and 2016. These studies explored the PubMed database within a 20-year period. However, our study explored SCD publications indexed in the Scopus database and extended beyond a 20-year period. Furthermore, no study has been found to have integrated systematic review and bibliometric visualization tools in providing a comprehensive and data-driven mapping of SCD research landscape in Africa.

This study aims to evaluate SCD research landscape in Africa and provide an updated research outcome on the documents published in Scopus database. This will provide information on the scientific output of SCD research in Africa and also identify the most influential contributors to SCD research in Africa and their impact.

Research questions:

1. What is the growth trend of SCD research in Africa?
2. Who are the most influential authors, institutions and countries that have contributed to SCD research in Africa?
3. What are the trending topics, major themes, and most used keywords in SCD research in Africa?
4. What pattern of collaboration exists in African SCD research among researchers and countries?

## 2.0. METHODOLOGY

This study seeks to fill a gap in the literature by integrating Systematic Literature review (SLR) and bibliometric analysis to evaluate SCD research landscape in Africa and provide an updated research outcome based on the documents published in Scopus database. The methodology we used for this study is described in Figure 1, which is a roadmap for the Systematic Literature Review and bibliometric analysis carried out. It began with the identification of what should be the focus of our study and choice of the database of scientific journals. This was followed by the collection of metadata using selected keyword strings that are related to our study. This was conducted in accordance with a predefined inclusion and exclusion criteria for the Systematic Literature Review based on the PRISMA 2020 statement. The retrieved dataset was manually screened to remove duplicates and irrelevant data which may give rise to misleading results and flawed interpretation. After which, the screened metadata was analyzed using VOSviewer and bibliometrix from which the graphical visuals of results were generated using biblioshiny, GraphPad prism, and datawrapper. Finally, the results were interpreted.

**Figure 1.**
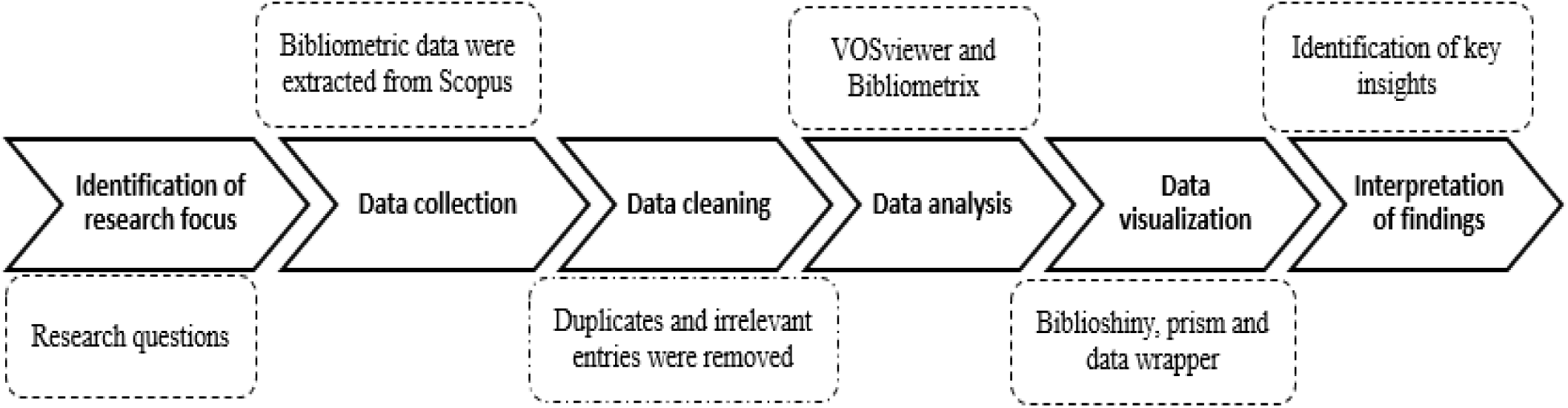
Study Flowchart.

### 2.1. Data Source

There are many databases (WoS, Scopus, Google Scholar, PubMed, Embase, Dimensions, Lens) that provide datasets for bibliometric analysis. Each of these databases has its unique characteristics. In comparison with other bibliometric databases, the Scopus database is better suited for bibliometric analysis as it is comprehensive and user friendly coupled with the fact that it is currently the largest curated database for abstracts and citations of peer reviewed literatures and as well as web sources (Agarwal *et al.,* 2015, Okoroiwu *et al.,* 2020). Also, it offers a comprehensive author and institution profiles and has a wide global and regional coverage of scientific journals (25,000), conference proceedings (6.5 million), and books (206,000) published by more than 5000 publishers worldwide (Baas *et al.,* 2020). Because of this, the Scopus database was chosen over others as shown in table 1 for this study.

**Table 1.**
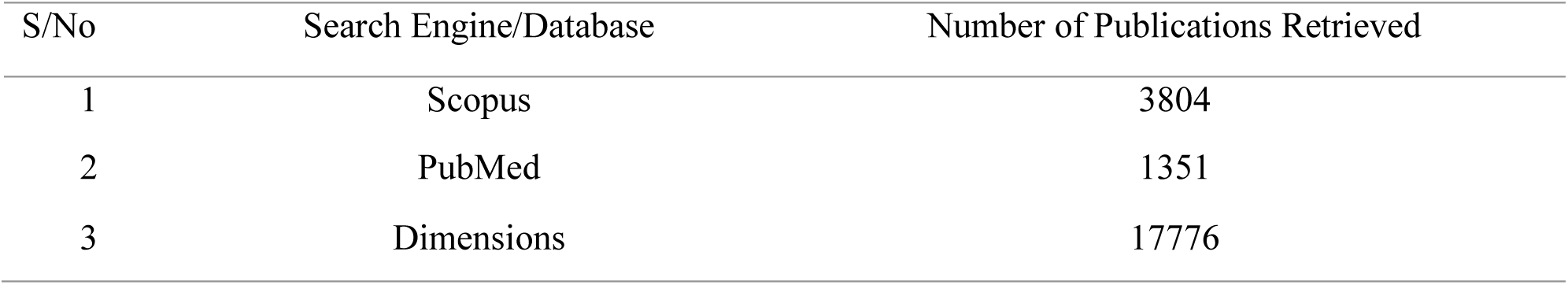
Number of Publications Related to Sickle Cell Disease in Africa Retrieved from Various Databases.

### 2.2. Data Collection

Data collection for this study was conducted in accordance with the Preferred Reporting Item for Systematic and Meta-Analysis (PRISMA) guidelines. The PRISMA method involves four (4) steps; identification, screening, eligibility, and inclusion/exclusion (Tawfik *et al*., 2019, Page *et al.,* 2021) as shown in figure 2.

**Figure 2:**
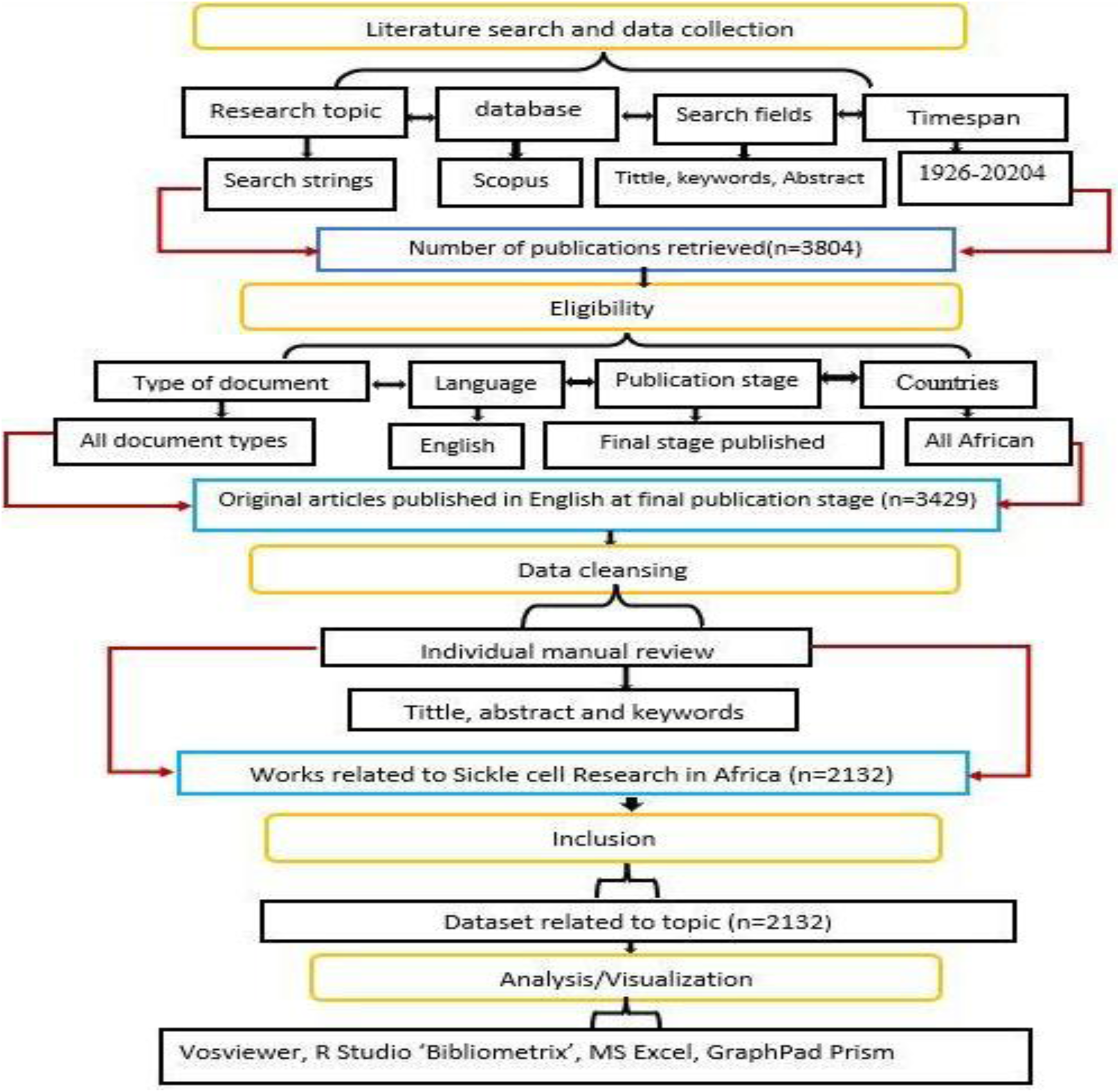
Flow Chart of our Search Strategy using PRISMA Guidelines.

#### 2.2.1. Identification

Data collection was conducted on 8^th^ August, 2024 at about 7:54 PM from the Scopus database which is known for its extensive academic publication coverage. The search strategy used in this study included specific search strings within the ‘Tittle-Abstract-Keywords’ field and they contain a combination of Boolean operators (OR and AND) and quotation marks (“…”). This is aimed at ensuring a comprehensive and relevant retrieval of datasets. The search strings used for this study are (“Sickle cell disease*” OR “Sickle cell anemia” OR “Sickle cell anaemia” OR “Drepanocytosis”) AND (“Africa”).

#### 2.2.2. Inclusion/Exclusion

We included all document types (original articles, conference papers, reviews, editorials, erratum, letter to the editor, short survey, etc.) that were published in English language and at the final publication stage from 1926 to 2024 in the records. While documents from non-English speaking countries and countries outside Africa were excluded. The records were then exported as a .csv file and a manual review of the retrieved dataset was then conducted to expunge documents not specifically related to sickle cell disease in Africa and duplicate copies.

### 2.3. Analysis and Visualization

For bibliometric analysis, Vosviewer version 1.6.20 (Leiden University, Leiden, Netherlands) (Van Eck & Waltman, 2017) was utilized for keywords co-occurrence and co-authorship. The R studio package ‘Bibliometrix’ was utilized for comprehensive bibliometric analysis. Graph pad prism version 8.0.2 and Datawrapper were used for data visualization.

## 3.0. RESULTS

### 3.1. An Overview of Research Statistics on Sickle Cell Disease Research in Africa

A total of 2132 publications on sickle cell disease research in Africa were identified in the Scopus database. This only concerns articles published in English at the final stage between 1926 to 2024. These 2132 publications have been authored by 7178 contributors across 728 sources (Figure 3). Among the 2132 publications, only 128 articles are with a single author.

**Figure 3.**
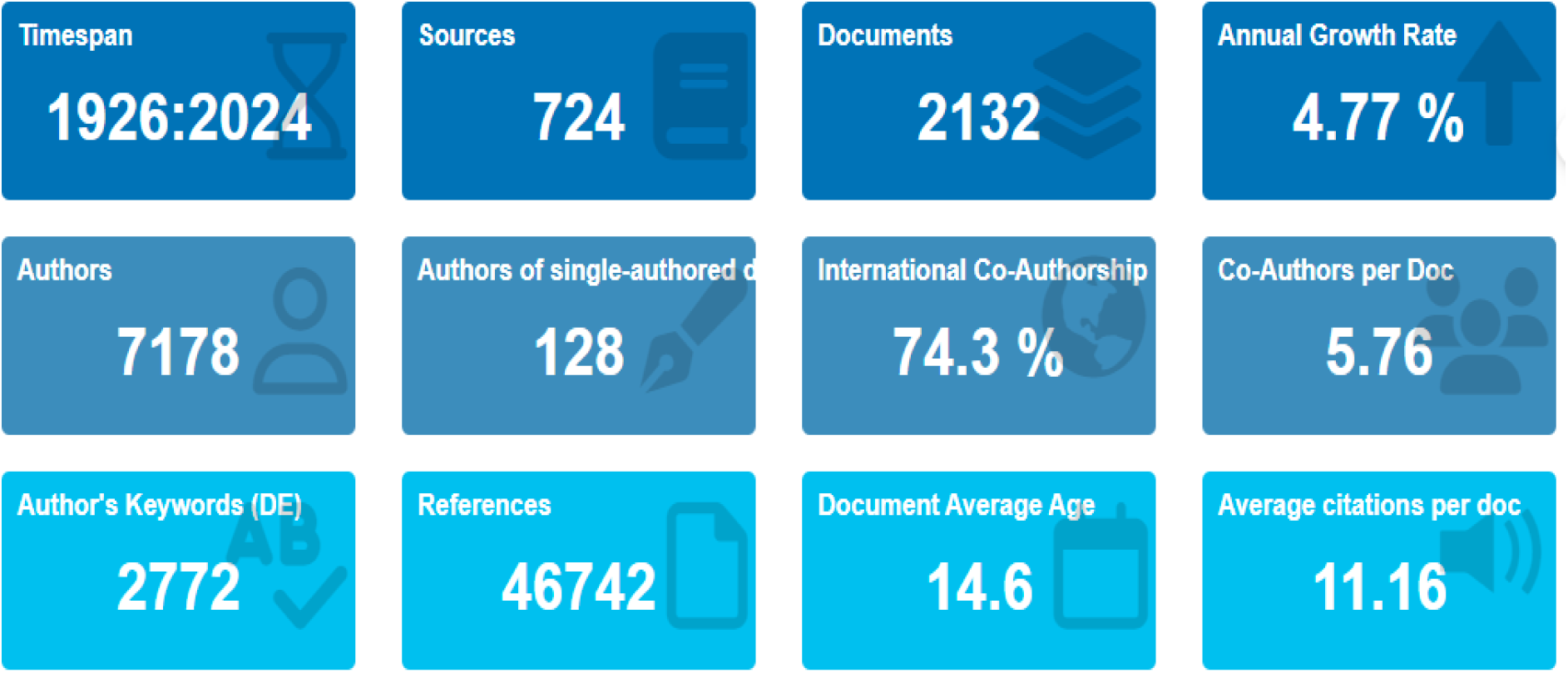
An Overview of Research Performance on Sickle Cell Disease in Africa Based on Scopus.

The number of articles published on sickle cell disease in Africa had consistently increased over time with an average annual growth rate of 4.77%. The international collaboration of 74.3% highlights a significant degree of international networking in research on sickle cell disease in Africa. The distribution of the various document types on SCD research in Africa showed that of the 2132 publications, 89.96% are original research articles (most abundant), 4.41% are review articles, 3.80% are letters, 1.22% are notes, 0.19% are short survey while 0.14% each are conference papers, editorials and erratum as shown in figure 4.

**Figure 4.**
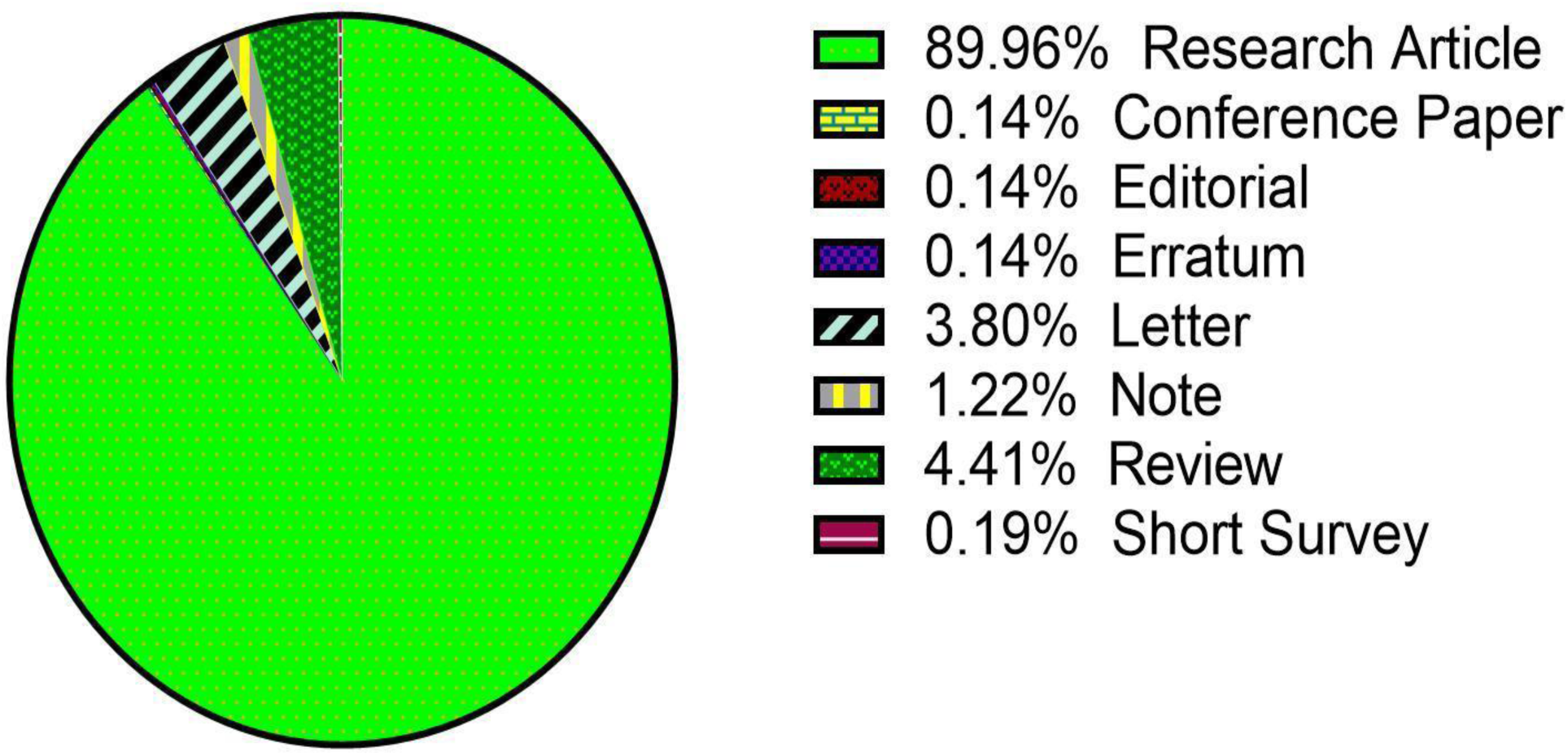
Distribution of Document Types on Sickle Cell Disease Research in Africa.

### 3.2. Publication Trends on Sickle Cell Disease in Africa

#### 3.2.1. Research Publication Trend

Figure 5 shows the annual publication output of articles on sickle cell disease in Africa. The first article on sickle cell disease in Africa was published in 1926 and was authored by Archibald, RG. Between 1927 to 1951, only 3 articles were published. This may be due to high publication fees associated with publishing in high-impact journals and also biases in global publishing systems. Furthermore, as of then, SCD was not widely recognized as a global health priority until 2006 when the World Health Organization (WHO) recognized it. A notable increase was observed from 2010 upward as this disease has drawn the attention of many more researchers since it has been recognized by WHO as a global health priority. The highest number of published articles on sickle cell disease in Africa was recorded in 2022, followed by 2020, 2019 and 2021 with 167, 130, 127 and 127 articles respectively. Figure 6 showed that the highest mean total citation per article were in 1970 (N=3) with 59.67 total citation per articles, followed by 1954 (N=1) and 1968 (N=1) with 34.00 and 33.00 total citations per article respectively. Also, the highest mean total citation per year were in 2011 (N= 63) with 1.85 mean total citation per year, followed by 2019 (N= 127), 2009 (N= 37), 2002 (N= 16), and 2017 with 1.70, 1.65, 1,52 and 1.44 mean total citations per year respectively.

**Figure 5.**
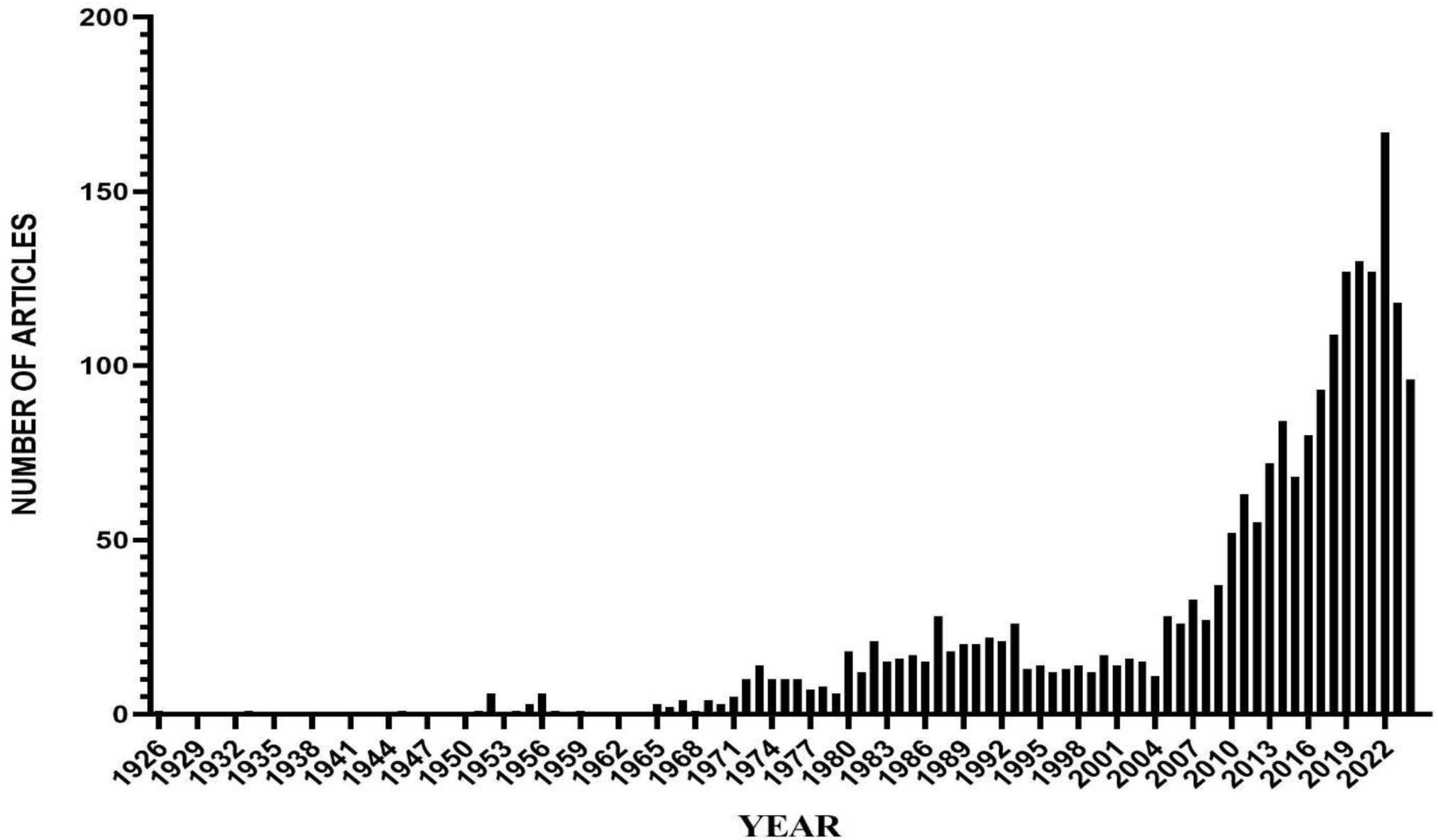
Annual Scientific Production of Articles on Sickle Cell Disease Research in Africa.

**Figure 6.**
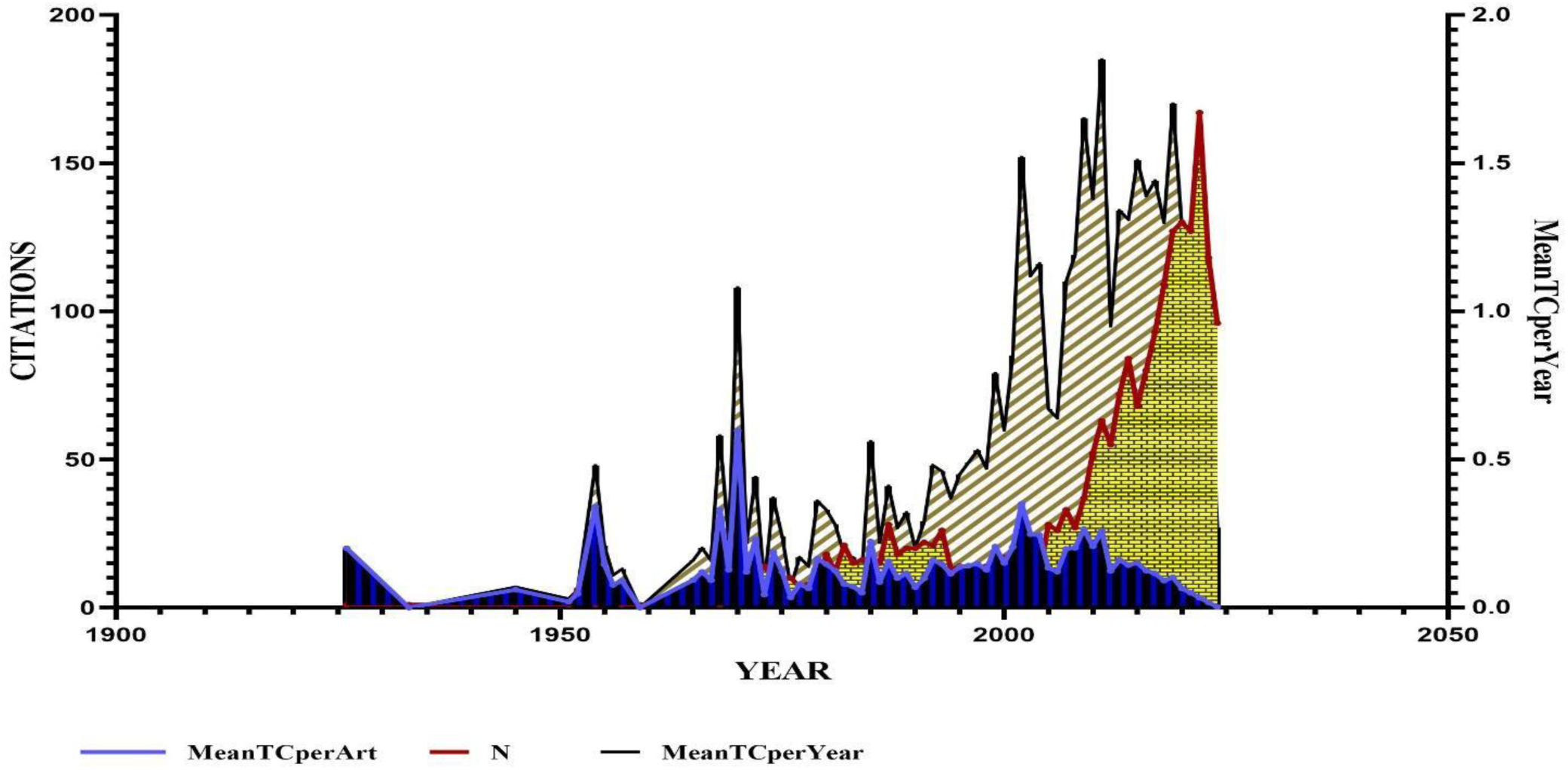
Total Citation per Year of Documents on Sickle Cell Disease Research in Africa.

#### 3.2.2. Geographic Distribution of Sickle Cell Disease Research in Africa

The scientific production map (figure 7a) shows the distribution of research output on sickle cell disease in Africa. In all, 89 countries have participated in research on SCD in Africa. Overall, Niger and Nigeria are the countries with the most frequency of publications on sickle cell disease in Africa as depicted by the darker regions on the map. Other countries with high publication frequency include Ghana, USA, Tanzania, Egypt and United Kingdom. Figure 7b shows countries research efforts on sickle cell disease in Africa overtime. As it can be visually observed, there is a consistent rise in research output on sickle cell disease in Africa overtime by the leading contributing countries. Niger and Nigeria have significantly contributed to the research output on sickle cell disease in Africa.

**Figure 7.**
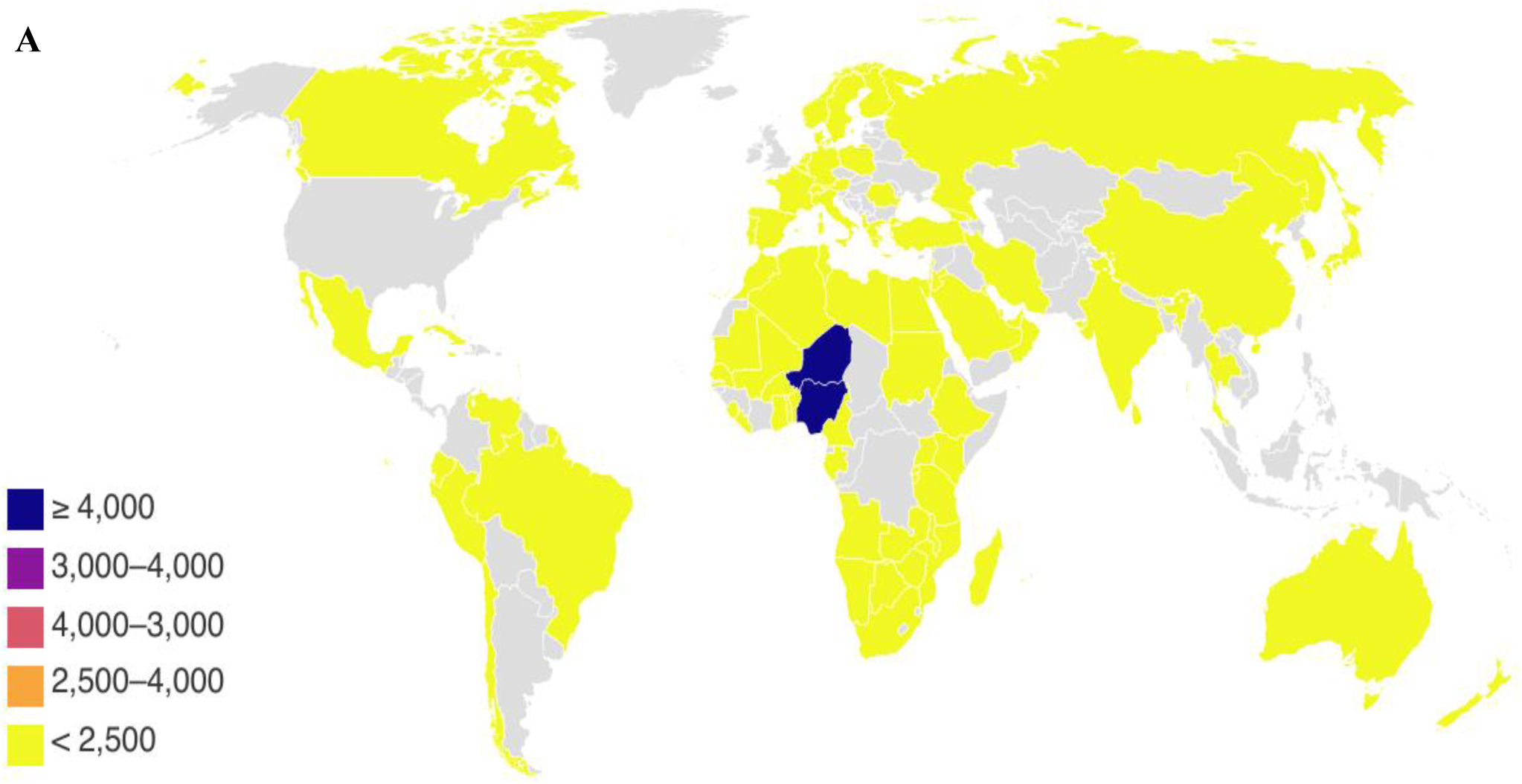
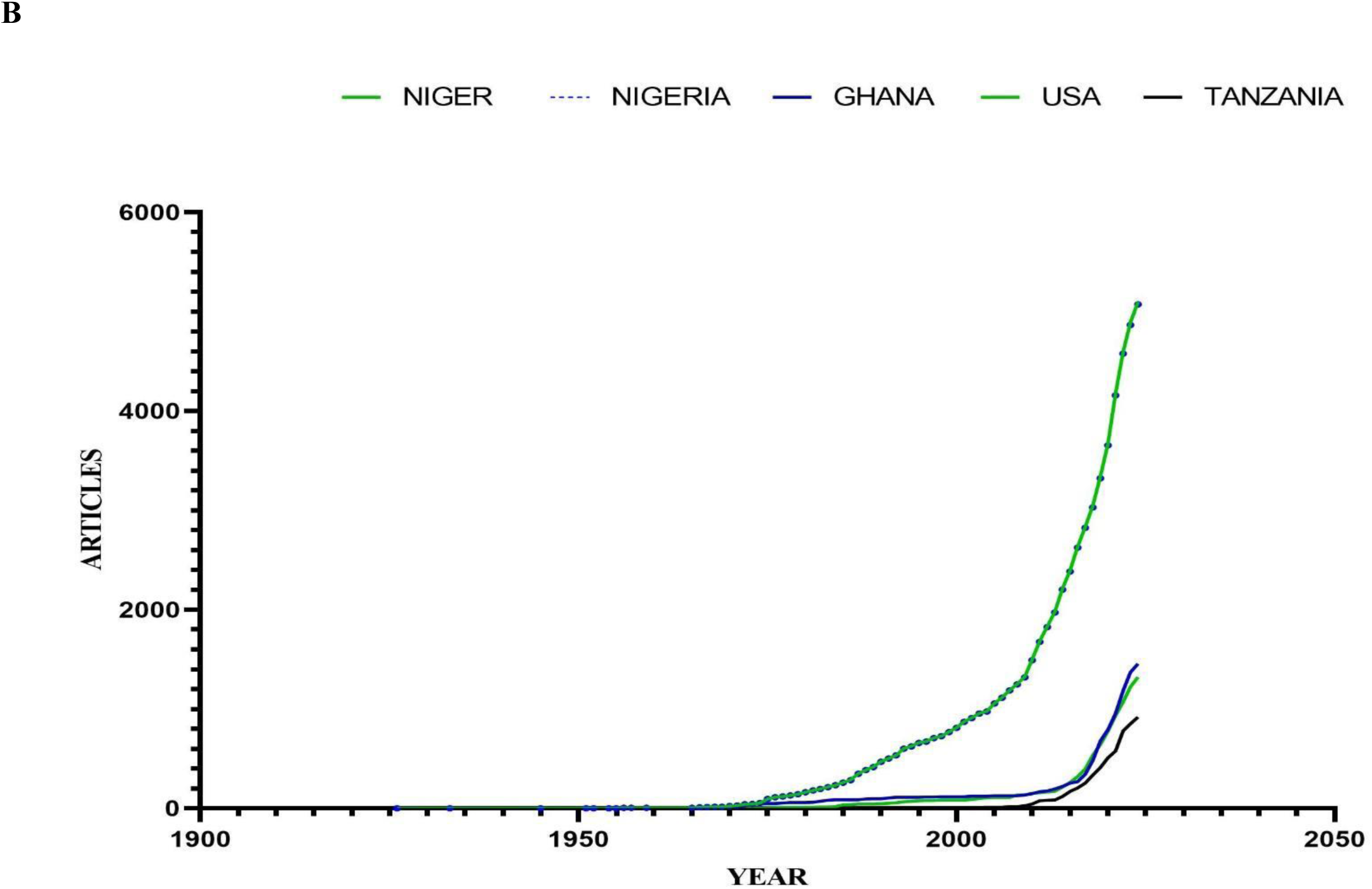
Countries Production on Sickle Cell Disease Research in Africa (A) Map of the World showing Countries that produced articles on Sickle Cell Research in Africa; (B) Countries Production Over Time on Sickle Cell Research in Africa

**Figure 8.**
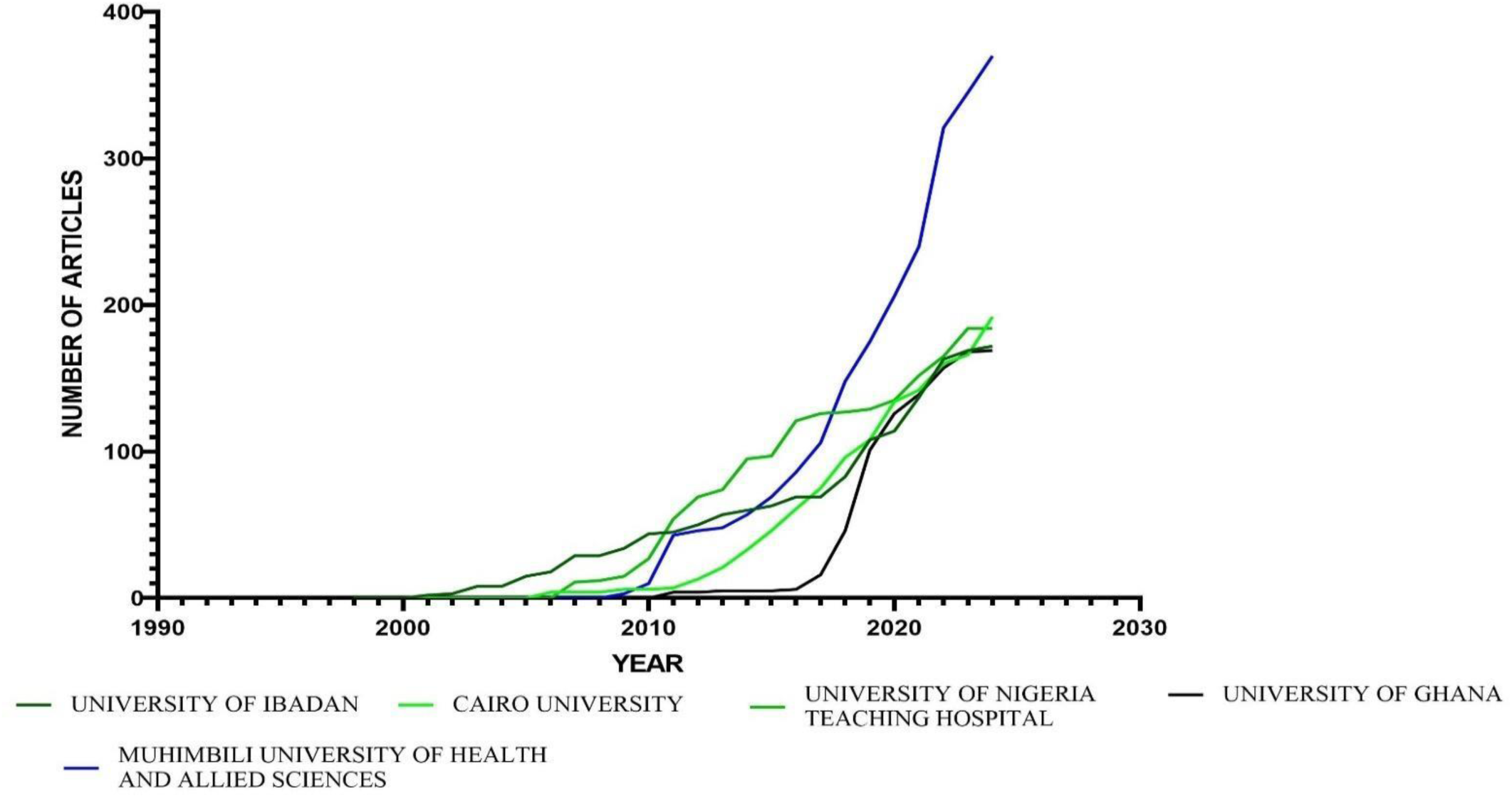
Affiliations’ Production Over time on Sickle Cell Disease Research in Africa.

**Figure 9.**
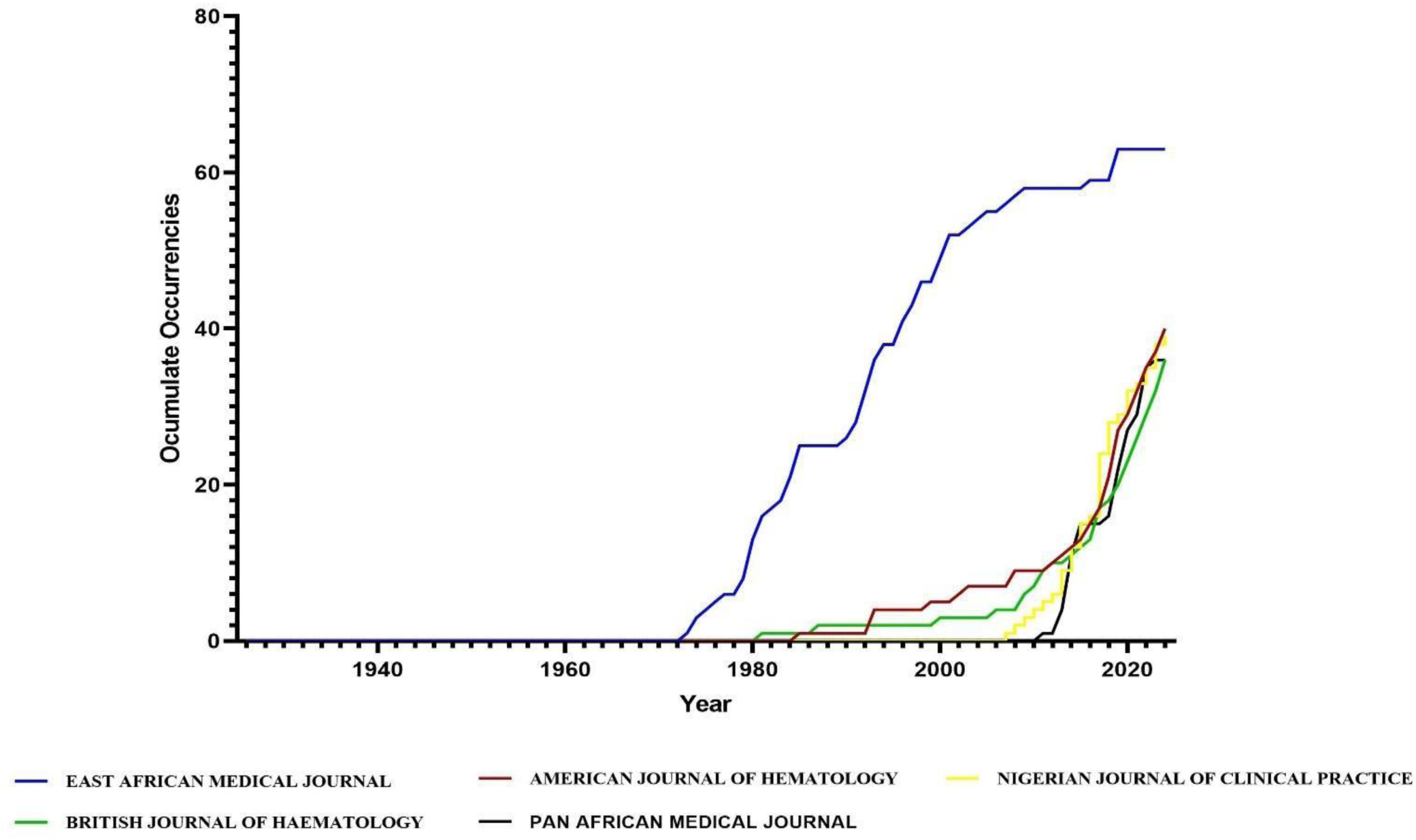
Source Production Overtime on Sickle Cell Disease Research in Africa.

### 3.3. Global Collaboration Network on Sickle Cell Disease Research in Africa

A significant collaboration on SCD research in Africa was observed from various countries in terms of single-country production (SCP) and multiple country production (MCP) (figure 10). This may be facilitated by shared research interests on sickle cell disease. Nigeria has more corresponding authors of publications on sickle cell disease in Africa, followed by Egypt, United State of America and Ghana. More research collaborations on sickle cell disease in Africa are reported among Nigeria, USA and Tanzania with more MCP whereas Morocco is the only country with SCP. In most of the corresponding authors’ countries, MCP is more than SCP.

**Figure 10.**
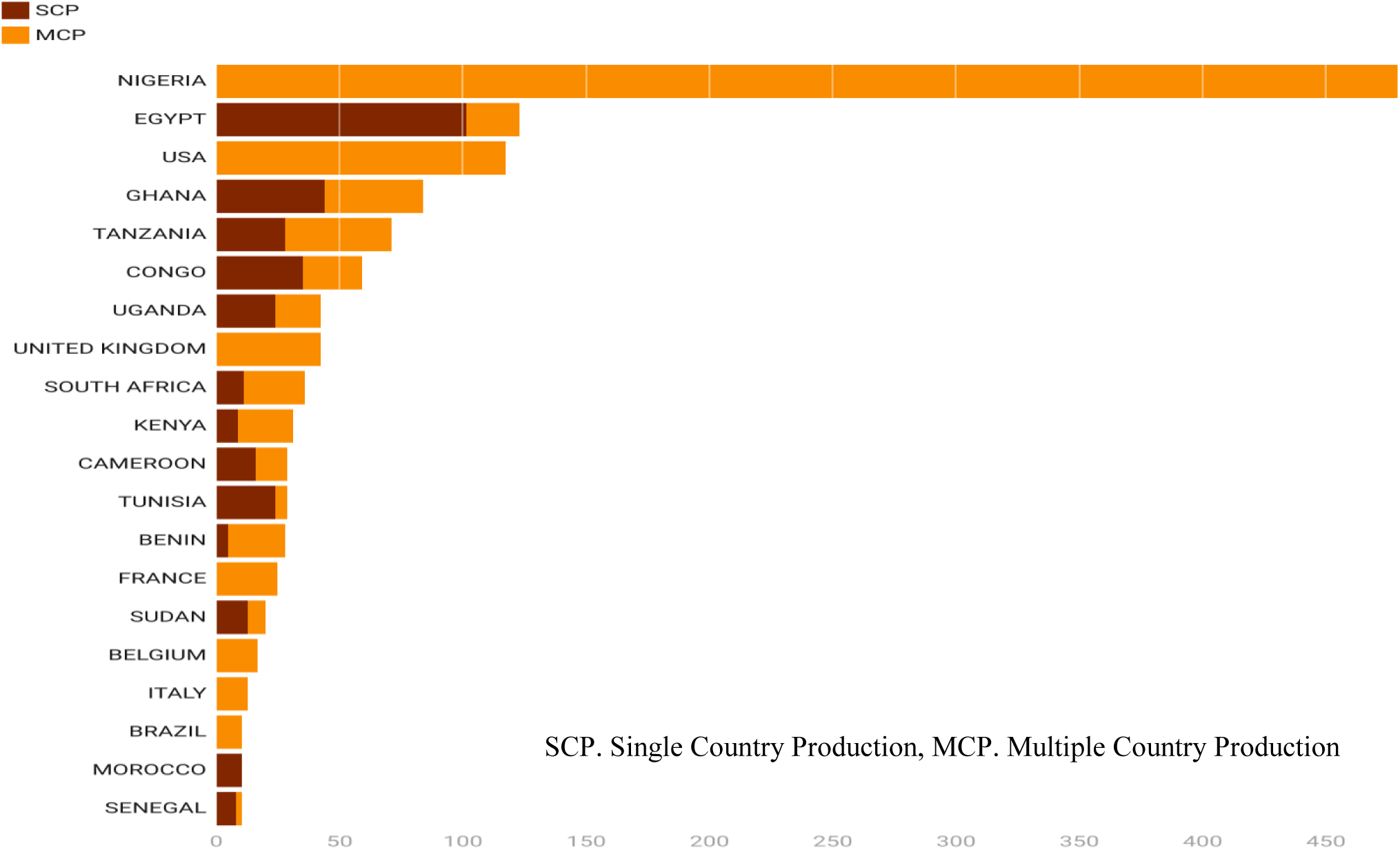
Corresponding Authors’ Countries Production on Sickle Cell Research in Africa.

Table 2 shows the top 10 countries that have the highest frequency of collaboration on sickle cell disease research in Africa. The highest frequency of collaboration is between Niger and Nigeria which occurs 1065 times.

**Table 2.** Top 10 Countries with the Highest Frequency of Collaboration in Sickle Cell Research in Africa.

| S/No | From | To | Frequency |
| --- | --- | --- | --- |
| 1 | Niger | Nigeria | **1065** |
| 2 | Niger | USA | 125 |
| 3 | Nigeria | USA | 125 |
| 4 | Niger | Benin | 72 |
| 5 | Nigeria | Benin | 72 |
| 6 | Nigeria | United Kingdom | 72 |
| 7 | Niger | United Kingdom | 72 |
| 8 | USA | United Kingdom | 70 |
| 9 | Ghana | USA | 63 |
| 10 | Tanzania | United Kingdom | 62 |

**Table 3.** Top 20 Countries Publication on Sickle Cell Disease Research in Africa Based on the Corresponding Author.

| S/No | COUNTRY | ARTICLES | SCP | MCP | % SCP TO TOTAL PUBLICATIONS | % MCP TO TOTAL PUBLICATIONS |
| --- | --- | --- | --- | --- | --- | --- |
| 1 | NIGERIA | 479 | 0 | 479 | 0 | 100 |
| 2 | EGYPT | 123 | 101 | 22 | 82.1 | 17.9 |
| 3 | USA | 117 | 0 | 117 | 0 | 100 |
| 4 | GHANA | 84 | 44 | 40 | 52.4 | 47.6 |
| 5 | TANZANIA | 71 | 28 | 43 | 39.4 | 60.6 |
| 6 | CONGO | 59 | 35 | 24 | 59.3 | 40.7 |
| 7 | UGANDA | 42 | 24 | 18 | 57.1 | 42.9 |
| 8 | UNITED KINGDOM | 42 | 0 | 42 | 0 | 100 |
| 9 | SOUTH AFRICA | 36 | 11 | 25 | 30.6 | 69.4 |
| 10 | KENYA | 31 | 9 | 22 | 29 | 71 |
| 11 | CAMEROON | 29 | 16 | 13 | 55.2 | 44.8 |
| 12 | TUNISIA | 29 | 24 | 5 | 82.8 | 17.2 |
| 13 | BENIN | 28 | 5 | 23 | 17.9 | 82.1 |
| 14 | FRANCE | 25 | 0 | 25 | 0 | 100 |
| 15 | SUDAN | 20 | 13 | 7 | 65 | 35 |
| 16 | BELGIUM | 17 | 0 | 17 | 0 | 100 |
| 17 | ITALY | 13 | 0 | 13 | 0 | 100 |
| 18 | BRAZIL | 10 | 0 | 10 | 0 | 100 |
| 19 | MOROCCO | 10 | 10 | 0 | 100 | 0 |
| 20 | SENEGAL | 10 | 8 | 2 | 80 | 20 |

### 3.4. Descriptive Statistics of Academic Journals and Most Cited Authors of Research on Sickle Cell Disease in Africa

Figure 11 shows the top 10 most productive institution on sickle cell research in Africa. Muhimbili University of Health and Allied Sciences in Tanzania is the most productive institution with 370 published articles, followed by Cairo University in Egypt with 197 articles, University of Nigeria Teaching Hospital with 184 articles, University of Ibadan in Nigeria with 172 articles and University of Ghana with 169 articles. Most of the most productive institutions are located in Nigeria. This is because Nigeria has the highest number of people with SCD globally. This high incidence created an urgent public health need which drives research on SCD in the country.

**Figure 11.**
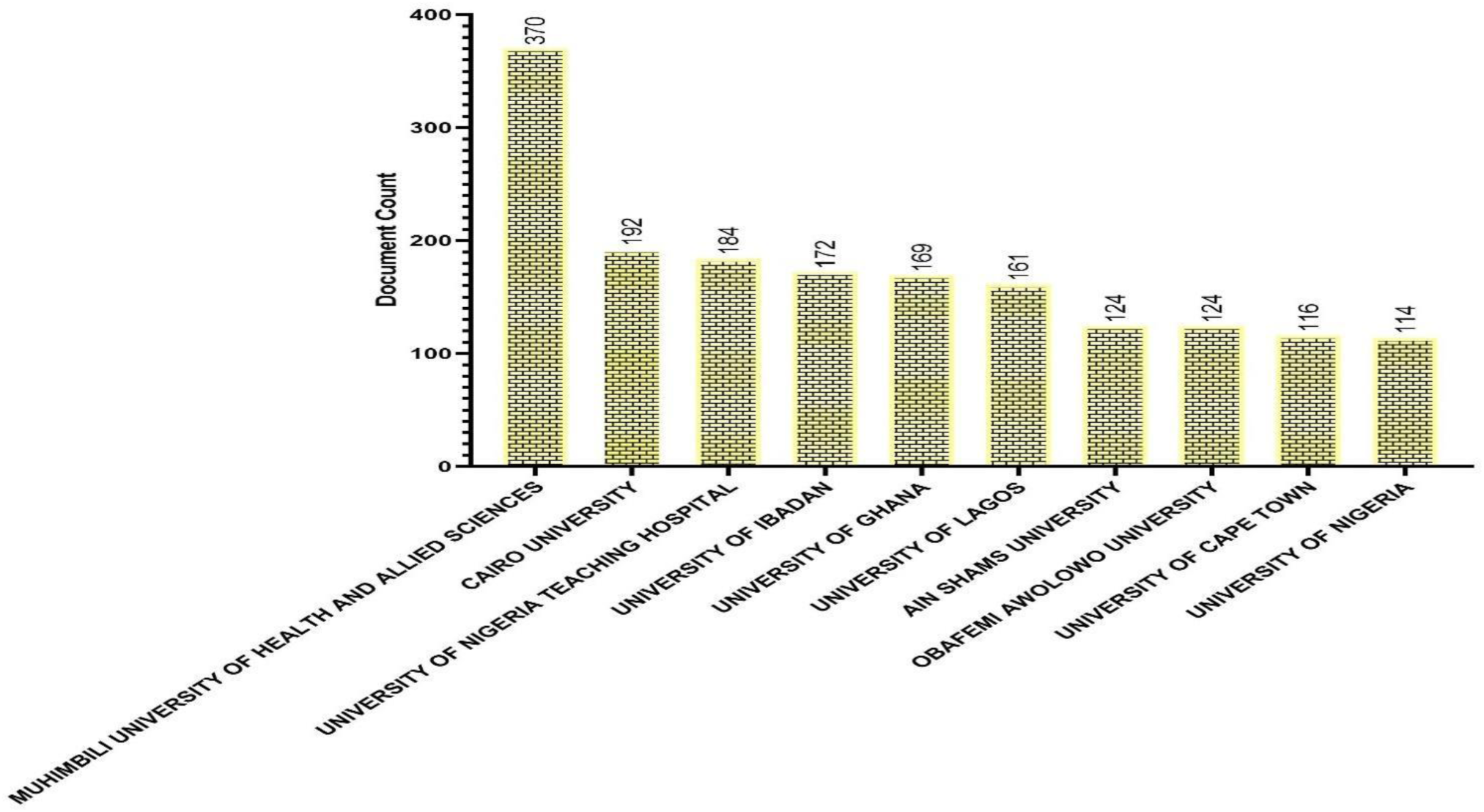
Top 20 Most Relevant Affiliation in Sickle Cell Disease Research in Africa.

**Figure 12.**
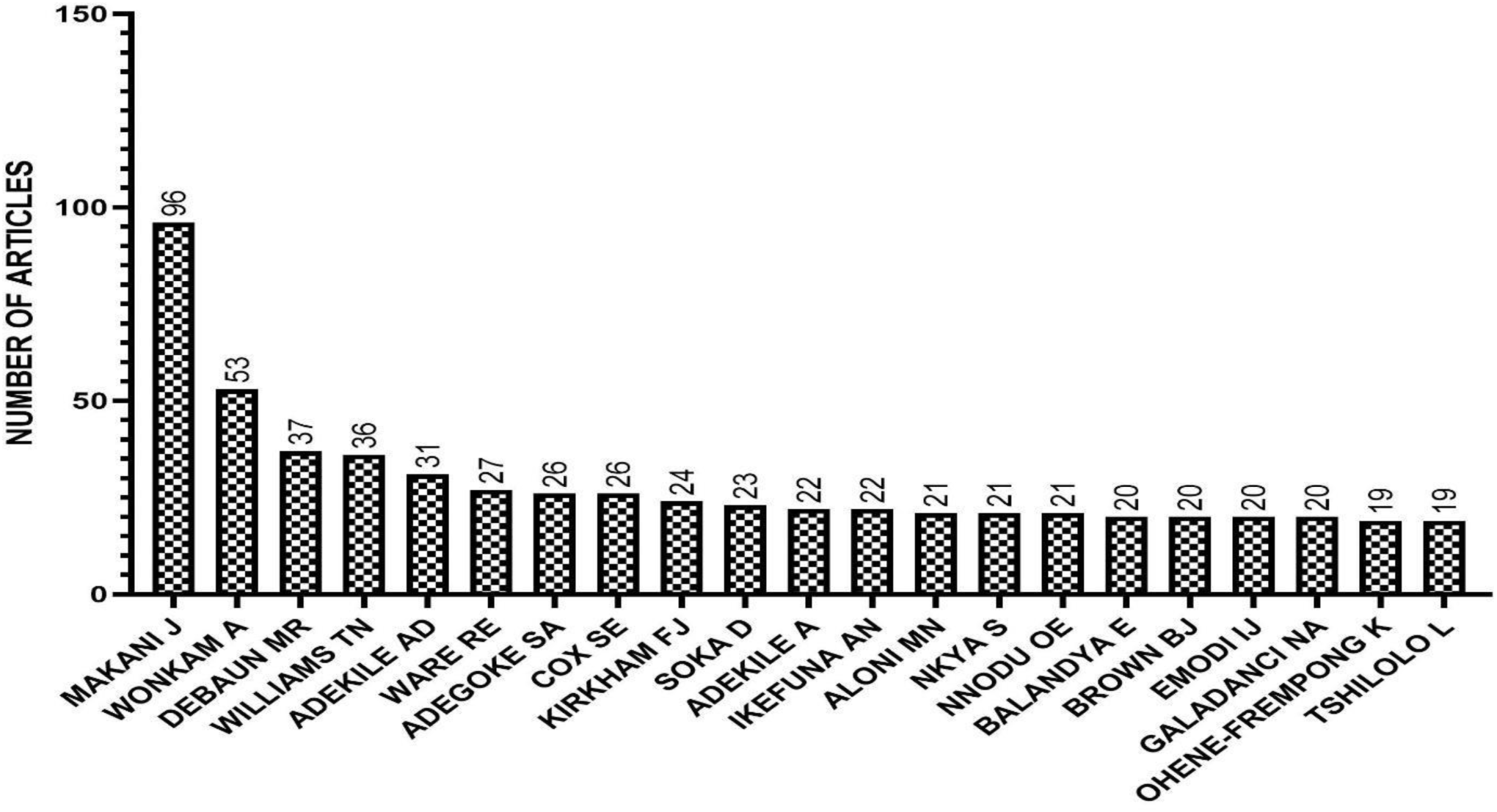
Top 20 Most Relevant Authors of Sickle Cell Research in Africa.

Table 4. shows the top 10 journals in which articles on sickle cell disease in Africa are published. The *American Journal of Hematology* which is a Q1 journal, is the most impactful journal with a Hirsch index (h-index) of 19 and a total citation of 887 from 40 publications followed by *Blood* which also is a Q1 journal with a h-index of 12 and a total of 792 citations from 14 publications. Table 5 shows the top 10 most cited articles on sickle cell disease research in Africa. The most cited article is a review article titled ‘Sickle Cell Disease in Africa: A Neglected Cause of Early Childhood Mortality’ by Gross *et al.,* (2011). This article has 445 citations and was published in *American Journal of Preventive Medicine*. Furthermore, the second most cited is also a review article titled ‘Hematologically and Genetically Distinct Forms of Sickle Cell Anemia in Africa: The Senegal Type and the Benin Type’ by Nagel *et al.,* (1985). This review has 236 citations and was pulished by *New England Journal of Medicine*.

**Table 4.** Top 10 Journals in which Sickle Cell Disease Research in Africa are published.

| S/No | Journals | H-index | Number of Publications | Total Citations | Q |
| --- | --- | --- | --- | --- | --- |
| 1 | American Journal of Hematology | 19 | 40 | 887 | Q1 |
| 2 | British Journal of Haematology | 13 | 36 | 543 | Q1 |
| 3 | Pediatric Blood and Cancer | 13 | 28 | 472 | Q1 |
| 4 | Plos One | 13 | 34 | 764 | Q2 |
| 5 | Blood | 12 | 14 | 792 | Q1 |
| 6 | Journal Of Tropical Pediatrics | 12 | 34 | 454 | Q2 |
| 7 | African Journal of Medicine and Medical Sciences | 11 | 29 | 275 | Q3 |
| 8 | East African Medical Journal | 11 | 63 | 375 | Q3 |
| 9 | Hemoglobin | 11 | 30 | 358 | Q3 |
| 10 | Tropical And Geographical Medicine | 11 | 27 | 255 | Q3 |

**Table 5.** Top 10 Influential Authors of Sickle Cell Disease Research in Africa.

| S/No. | Author | h_index | g_index | m_index | TC | NP | PY_start |
| --- | --- | --- | --- | --- | --- | --- | --- |
| 1 | Makani J | 23 | 43 | 1.095 | 2036 | 96 | 2004 |
| 2 | Williams TN | 19 | 36 | 1.056 | 2048 | 36 | 2007 |
| 3 | Wonkam A | 17 | 28 | 0.895 | 882 | 53 | 2006 |
| 4 | Cox SE | 16 | 26 | 1.067 | 928 | 26 | 2010 |
| 5 | Soka D | 16 | 23 | 1.143 | 777 | 23 | 2011 |
| 6 | Adekile AD | 14 | 22 | 0.35 | 526 | 31 | 1985 |
| 7 | Debaun MR | 14 | 22 | 1.4 | 513 | 37 | 2015 |
| 8 | Kirkham FJ | 13 | 24 | 0.813 | 654 | 24 | 2009 |
| 9 | Ware RE | 13 | 27 | 1.083 | 836 | 27 | 2013 |
| 10 | Brown BJ | 11 | 19 | 0.5 | 399 | 20 | 2003 |

**Table 5.**
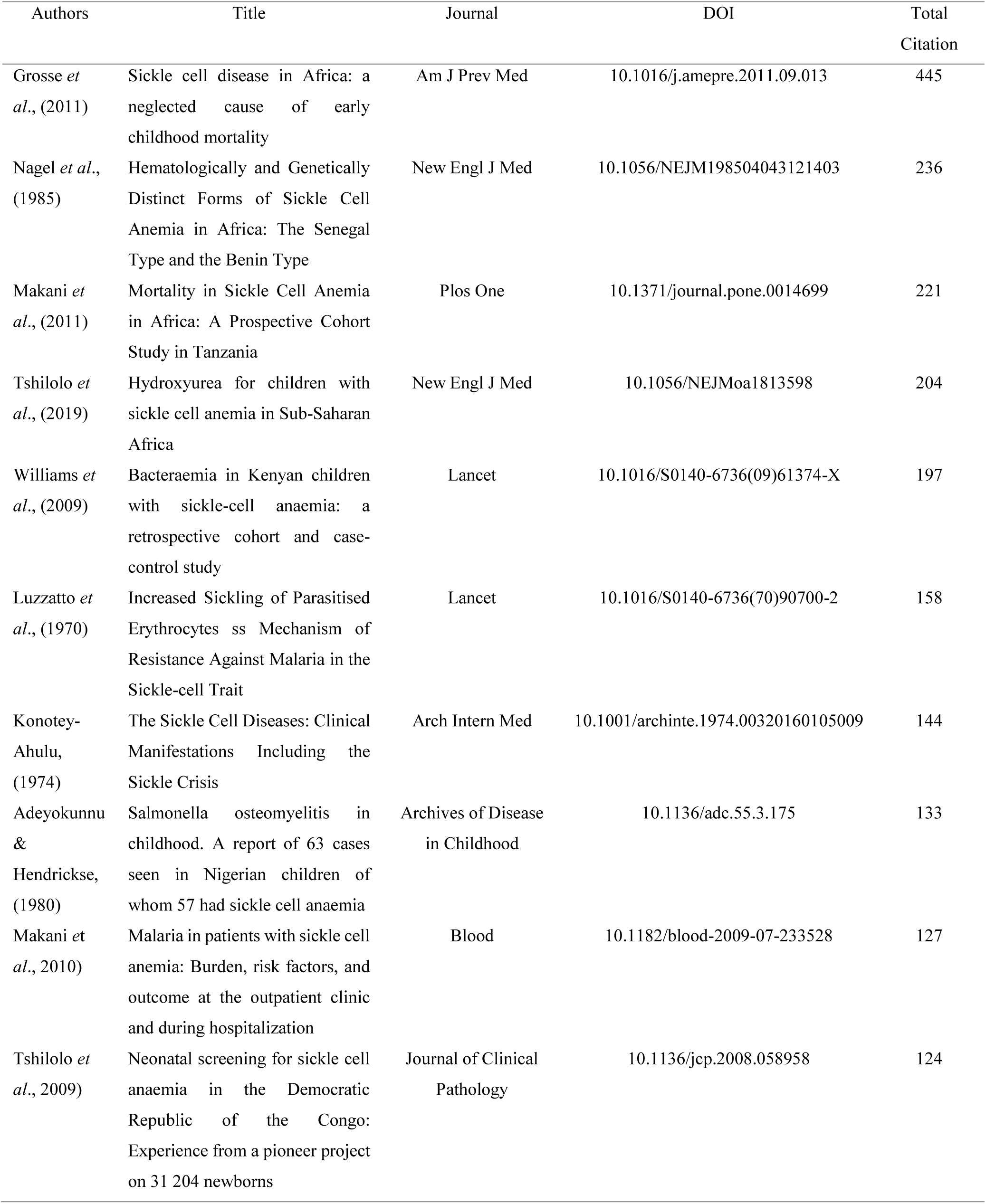
Most Cited Document on Sickle Cell Disease Research in Africa.

Table 5 shows the top 10 most prolific authors that have contributed to sickle cell disease research in Africa. Among the 7178 authors whose research on sickle cell disease in Africa were published, Williams T.N is the most cited author with a total citation of 2048 from 36 publications starting from 2007. This was followed by Makani J with a total citation of 2036 from 96 publications since 2004. Also, Cox, S.E had 928 total citations from 26 publications starting from 2010 and was followed by Wonkam, A and Ware, R.E with a total citation of 882 and 836 from 53, and 27 publications since 2006 and 2013 respectively. Other authors that have also contributed include; Soka D, Kirkham F.J, and Adekile, A.D with total citation of 777, 654, and 526 from 23, 24, and 37 publications since 2011, 2009 and 1985 respectively. Ranking the 7178 authors according to their impact on sickle cell disease research in Africa. Makani J. had the highest impact with an h-index of 23, followed by Williams T.N with 19 and Wonkam, A with 17.

Figure 13 shows the publication activity of various authors of articles on sickle cell disease research in Africa from 1985 to 2023. Each author’s publication activity is represented by circles along a horizontal line. The size of the circles indicates the number of published articles. The more significant the circle, the more the number of publications. The color intensity of each circle represents the number of total citations. The higher the number of total citations, the darker the color. As it can be seen from the production activities of top authors of publications on SCD in Africa, Adekile, A.D pioneered the initial production period in the authors’ productivity since 1985 with a total citation of 35 from 3 publications. This author had 56 total citations in 2017 from 4 publications as his highest. However, many other authors continued to produce several papers on SCD in Africa. In 2011, Makani, J had a total citation of 381 from 5 publications and in 2022, she had a publication frequency of 18 which is the highest frequency recorded in this study, similarly, that same year, Cox S.E also had a total citation of 381 from 5 publications. Wokam’s highest total citation of 264 was recorded in 2014 from 8 publications. In 2017, he had a publication frequency of 7. Debaun, M.R had a total citation of 135 from 3 publications in 2015 and in 2022, he had a publication frequency of 8 as the second highest recorded in this study. Williams, T.N recorded a total citation of 785 from 4 publications in 2011 (the darkest color) which is the highest total citation recorded in this study.

**Figure 13.**
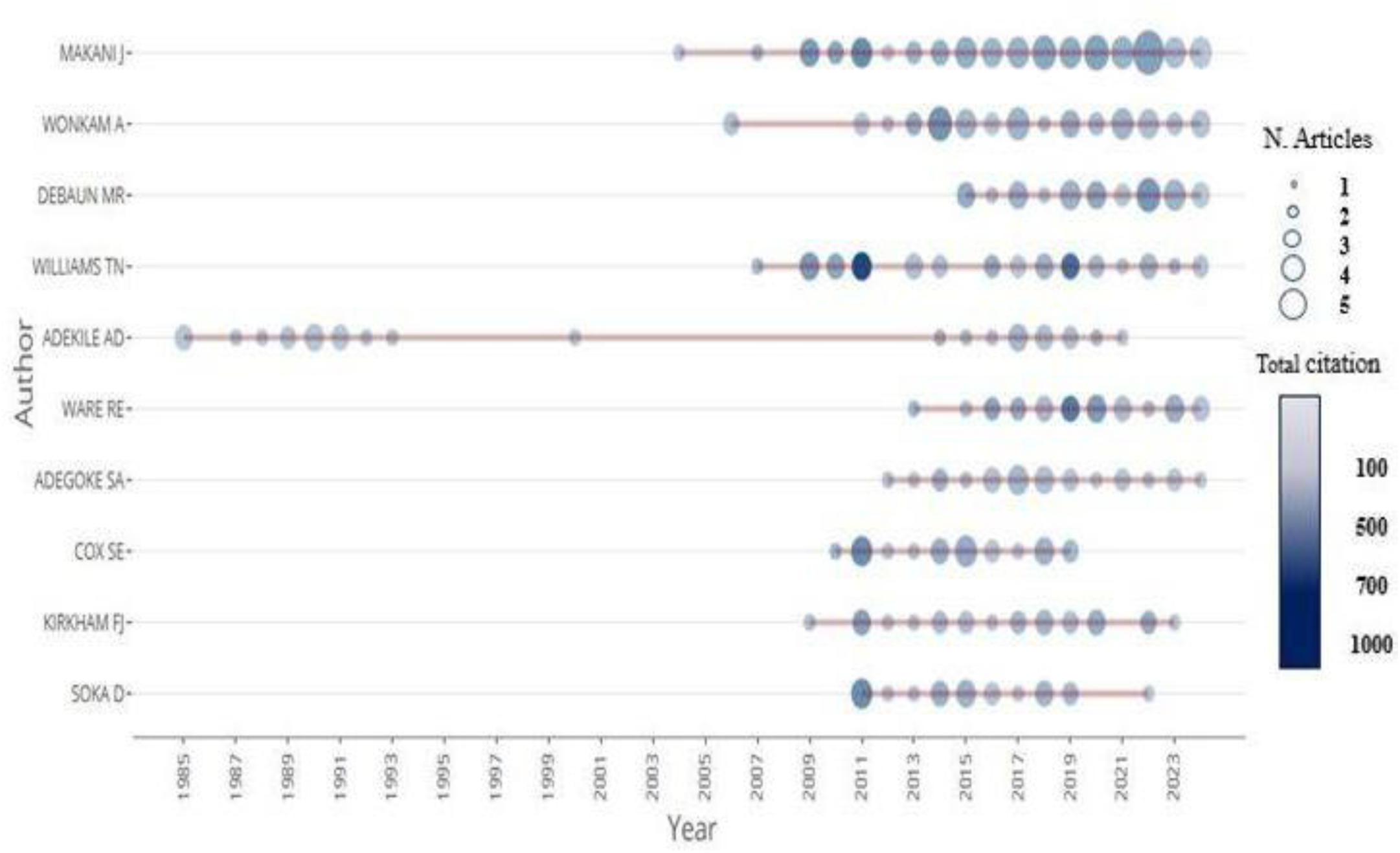
Top 10 Authors Production Overtime on Sickle Cell Disease Research in Africa.

Figure 15 depicts the top 20 most cited countries of sickle cell disease research in Africa. Nigeria is the most cited country with 3783 citations, followed by USA, Tanzania, United Kingdom and Kenya with 2536, 1189, 951 and 855 citations respectively.

**Figure 15.**
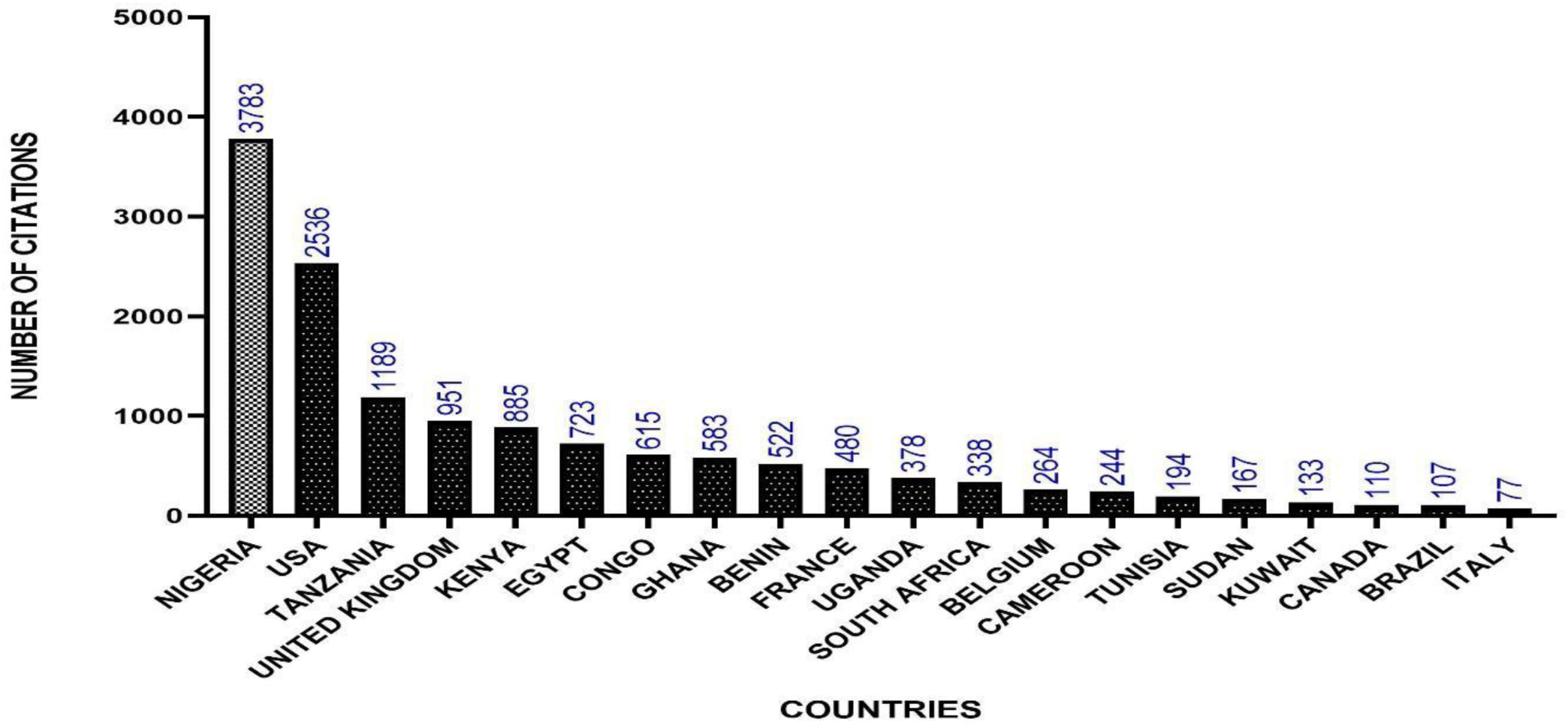
Top 20 Most Cited Countries on Sickle Cell Disease Research in Africa.

### 3.5. Emerging Themes of Research on Sickle Cell Disease in Africa

#### 3.5.1. Trending Topics of Research on SCD in Africa

Figure 16 shows the trending topics on sickle cell disease research in Africa. The term ‘erythrocyte’ was the most frequently occurring topic in 1994 with a frequency of 6. Thereafter, research focus shifted to magnesium, haplotypes and psychosocial in 2005, 2006 and 2007 with frequencies of 5, 7 and 5 respectively. In 2010, the terms sickle-cell anaemia, antisickling activity, and prophylaxis were the topics that trended with frequency of occurrences of 12, 8 and 6 respectively. Moving forward to 2011, prenatal diagnosis, and proteinuria trended with frequencies of 7 and 5 respectively. In 2012, the most commonly used terms were thalassemia, complications and nitric oxide which had frequency of occurrences of 10, 7 and 5 respectively. Forward to 2014, sickle-cell disease, sickle cell disorder and oxygen saturation were the terms that trended with frequencies of 20, 10 and 5 respectively. Child, malondialdehyde. And polymorphisms were the topics that trended in 2015 and had frequencies of 10, 6 and 5 respectively. In 2016, the trending topics were sickle cell anaemia (165), malaria (36), and pulmonary hypertension (18), while sickle cell anemia (214) and children (116) were the topics that trended in 2018. In 2019, sickle cell disease (572), stroke (36), and sub-Saharan Africa were the terms that trended. Moving forward to 2020, vaso-occlussive crisis (23), and fetal hemoglobin (16) were the topics that trended while hydroxyurea (59) and new born screening (24) were terms to have trended in 2021. In 2022, management and sickle cell disease, both with frequencies of 12, were the topics that trended. Lastly, in 2023, spleen was the trending term with frequency of occurrence of 8.

**Figure 16.**
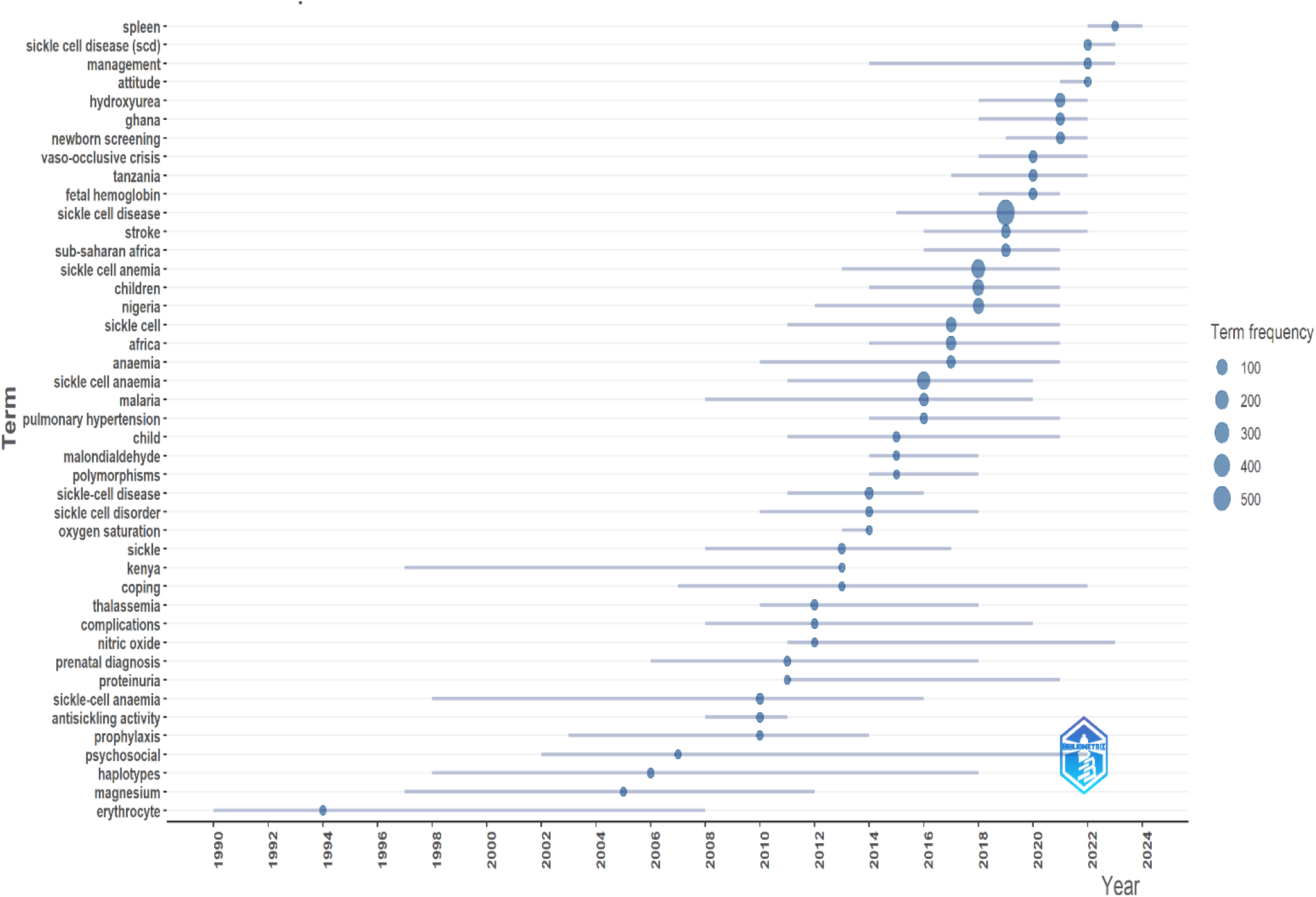
Trending Topics on Sickle Cell Disease Research in Africa.

#### 3.5.2. Thematic Map of Authors’ Keywords used in SCD in Africa

Figure 17 is a thematic map based on author’s keywords used in SCD research in Africa. It shows the relationship between different keywords used in SCD research in Africa. There are four quadrants; the basic theme, located at the bottom right quadrant, consisted of keywords such as ‘sickle cell disease’, ‘sickle cell anemia’ and ‘anti-sickling ‘. These themes have low density but high centrality. The motor theme, positioned at the upper right quadrant, represents the current research focus on SCD in Africa. These themes have high density with low centrality. The keyword Sickle cell disease (scd) was found here. Located at the bottom left quadrant is the emerging theme which represents areas of growing research interest. Keywords such as ‘fetal hemoglobin’, and ‘oxidative stress’ are found in this region. Located at the top left quadrant is the niche theme which represents specialized or less explored areas of research on SCD in Africa. Keywords such as ‘sickle cell trait’, ‘priapism’ and ‘thalassemia’ are found in this region.

**Figure 17.**
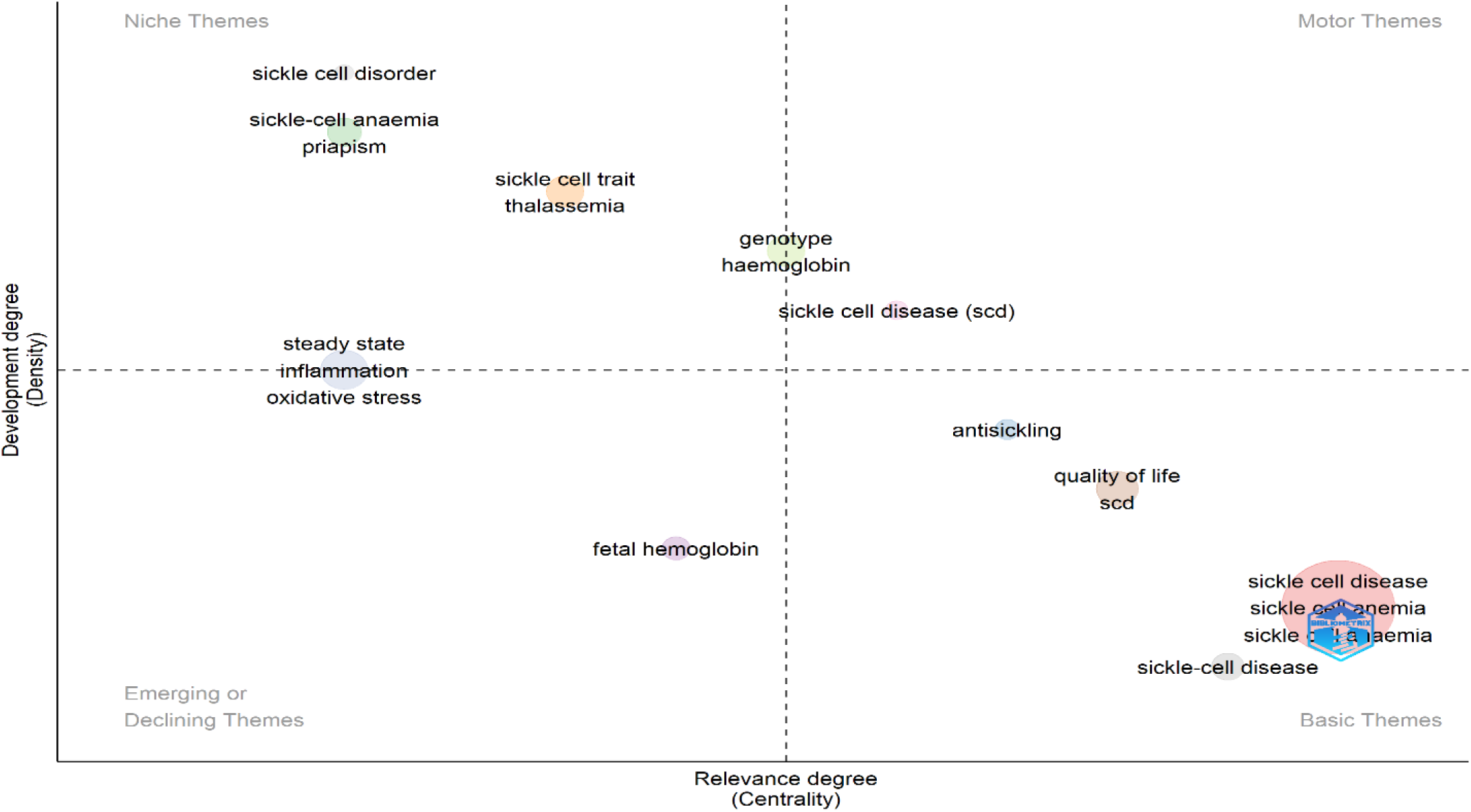
Thematic Map Based on Authors’ Keywords in Sickle Cell Disease in Africa.

#### 3.5.3. Thematic Evolution of Authors’ Keywords used in SCD Research in Africa

Thematic evolution map depicts the history of research themes and how these themes evolved over time (Nasir *et al.,* 2020). Figure 18 shows the thematic evolution and relationship between author’s keywords used in SCD research in Africa. The segmentation of time is based on the subjective judgement of the authors. These time segments have been grouped into three (3) periods of analysis which clearly show a shift in research focus on SCD in Africa. The period between 1926-2010 was dominated by topics such as ‘sickle cell disease’, ‘sickle cell anemia’ and ‘anti-sickling’. The period between 2011-2019 saw a surge in a broader range of terms such as ‘epidemiology’, ‘inflammation’, ‘anti-sickling’ ‘quality of life’, ‘oxidative stress’ and ‘vasculopathy’. Topics related to sickle cell evolved into hemoglobinopathies, sickle cell disease and sickle cell trait. Furthermore, parts of sickle cell anaemia evolved into sickle cell disease and the other parts into sickle-cell anaemia and steady state. Also, sickle cell disease evolved into β-thalassemia, L-arginine, anti-sickling, blood transfusion, chronic kidney disease, sickle cell anemia and fetal hemoglobin from 2020-2024. During this period, oxidative stress and sickle cell trait were introduced and both eventually evolved into sickle cell disease and sickle cell anemia from 2020-2024.

**Figure 18.**
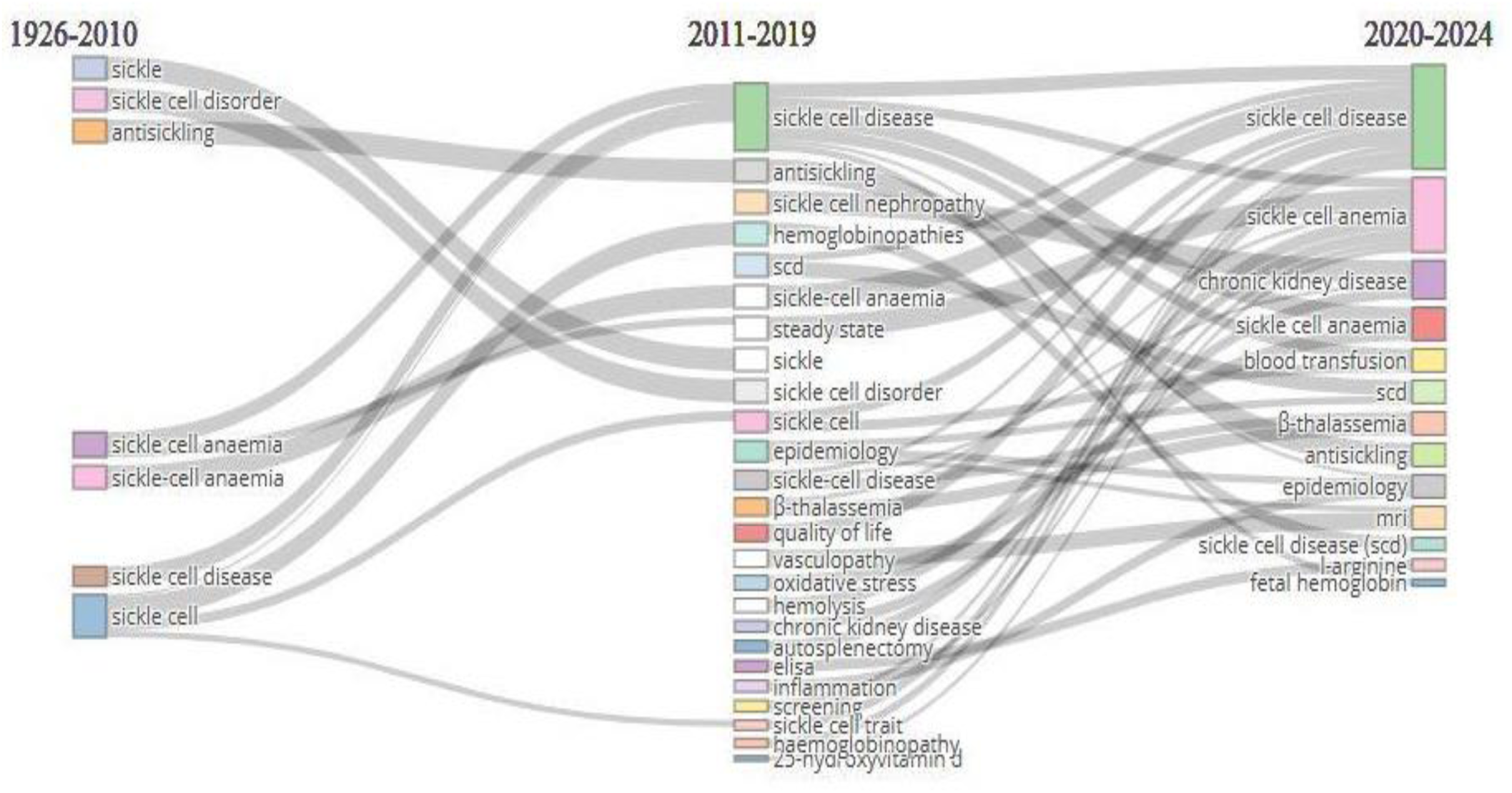
Thematic Evolution of Authors Keywords on Sickle Cell Disease Research in Africa.

### 3.6. Co-occurrence Network Analysis of Keywords

Keywords co-occurrence analysis is an effective tool that provides insights into knowledge structures and research trends in a particular field (Borgohain *et al.,* 2022; Rojas-Sánchez *et al.,* 2023). It facilitates the identification of the principal research areas. A graphical depiction of keywords provides insights into the various topics covered and how these topics interrelate (Belu & Marinoiu, 2025). Keyword co-occurrence network map consists of nodes and lines. Each node represents a keyword while the size of the node designates the number of documents. The line connecting two nodes represents co-occurrence between the two keywords. The shorter the line, the stronger the co-occurrence and vice versa. Color similarity signifies close relationship. We used the VOSviewer software to conduct a keyword co-occurrence analysis of all-keywords, authors’ keywords and indexed keywords that have been used in SCD research in Africa.

#### 3.6.1. Co-occurrence of All-keywords

In this study, the co-occurrence of all-keywords was evaluated. A total of 99000 keywords were identified. Taking the minimum number of occurrences of a keyword to be 5, 1612 keywords out of the total keywords met this threshold and 1290 keywords were used to construct the co-occurrence network map as shown in figure 19. The keywords were divided into 9 clusters. Cluster 1(red color) contains 323 keywords. The keyword ‘blood transfusion’, has the highest link strength and occurrence. Cluster 2 (green color) has 186 keywords with ‘blood’ having the highest link strength and occurrence. Cluster 3 (deep blue) consists of 158 keywords. The term ‘sickle cell anemia’ has the highest link strength and occurrence. Cluster 4 (yellow color) is made up of 146 keywords. ‘Genetics’ is the keyword with the highest link strength and occurrence. Cluster 5 (violet color) contains 138 keywords with ‘hemoglobin’ as the keyword with the highest link strength and occurrence. Cluster 6 (shallow blue) has 116 keywords. The term ‘human cell’ has the highest link strength and occurrence. Cluster 7 (orange color) consists of 86 keywords with ‘blood and hemopoietic system’ having the highest link strength and occurrence. Cluster 8 (brown color) is made up of 72 keywords. Children have the highest strength and occurrence. And lastly cluster 9 (purple color) has 65 keywords with prevalence as the keyword with the highest link strength and occurrence.

**Figure 19.**
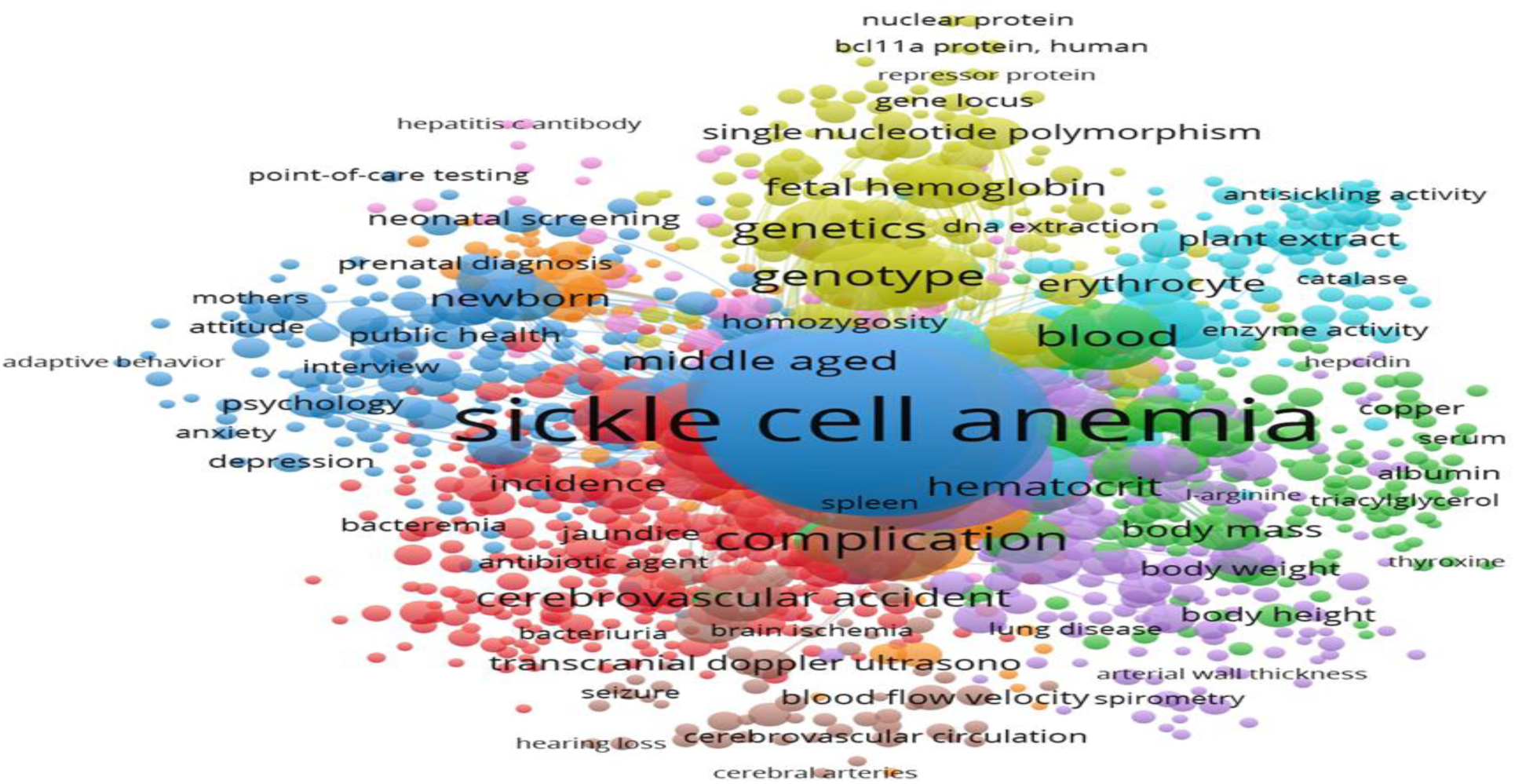
Co-occurrence network of All-Keywords used in Sickle Cell Disease Research in Africa.

#### 3.6.2. Co-occurrence of Authors’ Keywords

The co-occurrence analysis of authors’ keywords provides an insight into the most frequently used keywords by the authors and how these keywords interrelate. In this analysis, a total of 2773 keywords were identified. Taking the minimum number of occurrences of a keyword to be 5, it was found that 151 keywords meet the threshold and 131 keywords were used to construct the co-occurrence network map of the authors’ keywords as shown in figure 20. These connected keywords are divided into 13 clusters, each with a definite color. Cluster 1 (red color) contains 22 keywords. Some of the keywords include ‘ant sickling’, ‘antioxidant’, ‘sickle cell anemia’, ‘steady state’, ‘oxidative stress’. ‘Sickle cell anemia’ shows the highest strength and occurrence. Cluster 2(green color) has 19 keywords. Some of them are ‘hemoglobinopathy’, ‘foetal haemoglobin’, ‘new born screening’, ‘alpha thalassemia’, ‘genetic counselling’. In this cluster, ‘hemoglobinopathy’ is the keyword with the highest link strength while ‘new born screening’ has the highest occurrence. Cluster 3 (blue) contains 14 keywords including ‘sickle cell disease’, ‘quality of life’, ‘genotype’, and ‘psychosocial’. The keyword ‘sickle cell disease’ shows the highest link strength and occurrence. Cluster 4 (yellow color) is made up of 14 keywords. Some of which are ‘fetal hemoglobin’, ‘vaso-occlussive crises’, ‘hematology’, and ‘nephropathy’ with ‘hemoglobinopathy’ as the keyword with the highest link strength and occurrence. Cluster 5(mauve color) contains 14 keywords, some of which are ‘diagnosis’, ‘vasculopathy’, ‘stroke’,’ retinopathy’ and ‘prevalence’. The keyword ‘stroke’ has the highest link strength and occurrence followed by ‘prevalence’ and ‘management’. Cluster 6(light blue) has 9 keywords. Some of them are ‘cytokines’, ‘hydroxyurea’, ‘nutritional status’. ‘Hydroxyurea’ is the keyword with the highest link strength and occurrence. Cluster 7 (orange) consists of 9 keywords. Among them are l-arginine, anaemia, nitric oxide, genetics, pulmonary hypertension. The keyword that has the highest link strength and occurrence is ‘pulmonary hypertension’. Cluster 8(brown color) has 8 keywords. Some of them are ‘glomerular filtration rate’, ‘sickle cell nephropathy’, ‘micro albuminuria’. The keyword ‘children’ has the highest link strength and occurrence. Cluster 9(purple color) consists of 7 keywords including antioxidants, disease severity, haemoglobinopathies, inflammation, sickle cell anaemia. Among these keywords, sickle cell anaemia has the highest link strength and occurrence. Cluster 10 (light red) contains 6 keywords; ‘bacterial infection’, ‘hematological parameter’, ‘proguanil’, ‘sickle-cell disease’, ‘prophylaxis’ and ‘child’ with sickle-cell disease having the highest link strength and occurrence. Cluster 11(light orange) is made up of 5 keywords; ‘alloimmunization’, ‘blood transfusion’, ‘hepatitis c’, ‘iron overload’ and ‘serum ferritin’. ‘Blood transfusion’ has the highest link strength and occurrence. Cluster 12(pink) consists of 3 keywords; acute chest syndrome, cholelithiasis, and ultrasonography. Cluster 13(light green) contains ‘antisickling activity’ as the only keyword.

**Figure 20.**
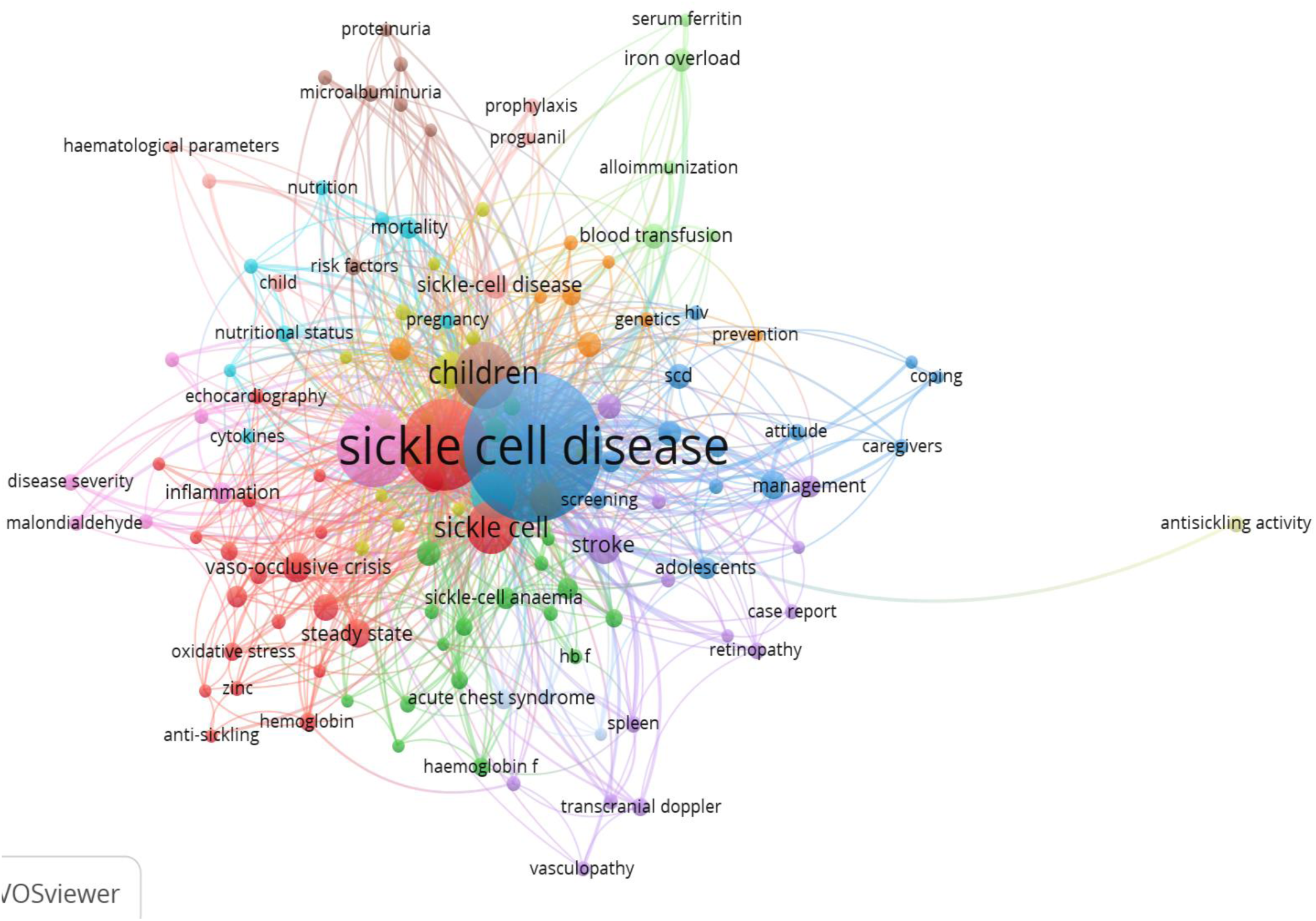
Co-occurrence Network of Authors’ Keywords used in Sickle Cell Disease Research in Africa.

#### 3.6.3. Co-occurrence of Indexed Keywords

The co-occurrence analysis of indexed keywords reveals how frequent certain keywords appears together in an indexing database. From the analysis, a total of 8001 indexed keywords were identified. Maintaining the minimum threshold of co-occurrence as 5, 1524 indexed keywords meet this threshold. Of this 1524 indexed keywords, 1061 were used to construct the co-occurrence network map as shown in figure 21. The co-occurrence network analysis of the indexed keywords identified 11 distinct clusters. Cluster 1 in red color, comprised of 277 indexed keywords with ‘blood transfusion’ as the most occurring indexed keyword among others such as ‘hydroxyurea’, ‘acute chest syndrome’, ‘blood count’, ‘antibiotic prophylaxis’ and ‘acute pain’. Cluster 2(green) contained 157 indexed keywords. Some of them are ‘acute kidney failure’, ‘alkaline phosphatase’, ‘bilirubin’, ‘creatinine’, ‘glomerulus filtration rate’. ‘Steady state’ is the most occurring indexed keyword. Cluster 3(blue) comprised of 126 indexed keywords. beta globin, cell adhesion molecules, beta-thalassemia, fetal hemoglobin. The most occurring indexed keyword is ‘genotype’. Cluster 4 (yellow) consisted of 104 indexed keywords including biomarkers, carnitine, lactate dehydrogenase, hemolysis. In this cluster, ‘hemoglobin’ is the most occurring indexed keyword. Cluster 5 (violet color) contained 98 indexed keywords. some of them are; ‘antianemic agent’, ‘antisickling agent’, ‘blood rheology’, ‘antioxidants’, ‘blood viscosity’. The term ‘hemoglobin s’ is the most occurring. Cluster 6 (light blue) has 85 indexed keywords with ‘sickle cell anemia’ as th most occurring indexed keyword. Cluster 7 (orange color) had 85 indexed keywords. Among them, ‘human’ is the most occurring indexed keyword. Cluster 8 (brown color) had 66 indexed keywords. some of them are; C-reactive proteins, cytokines, inflammation, interleukin 6, alloantibody. ‘Leukocyte count’ is the most occurring indexed keyword. Cluster 9 (purple color) contained 53 indexed keywords. Cluster 10 (light red) had 9 indexed keywords and cluster 11 (light green) had ‘spleen size’ as the only indexed keyword.

**Figure 21.**
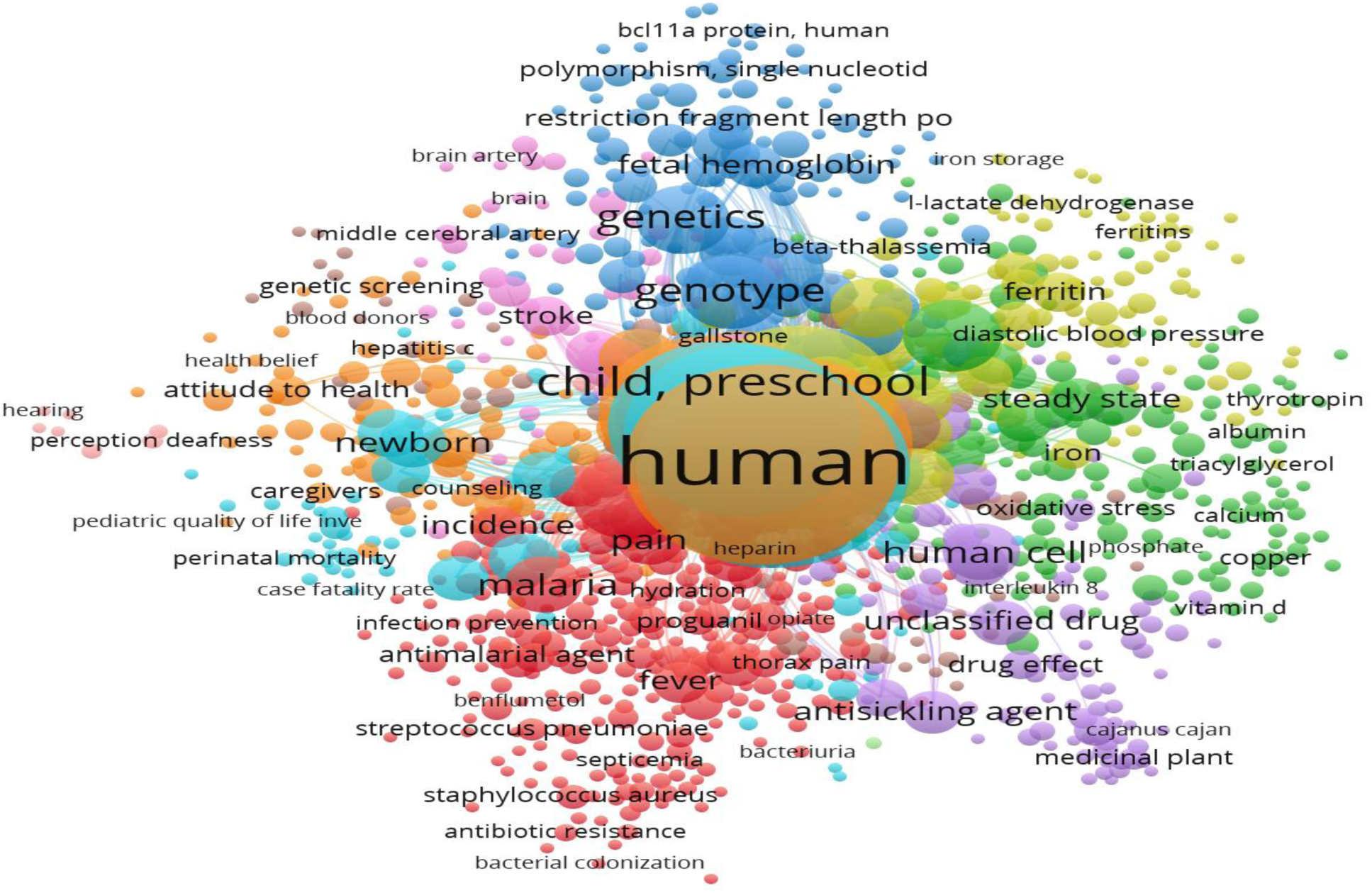
Co-occurrence Network Map of Indexed Key Words in Sickle Cell Disease Research in Africa.

### 3.7. Network Analysis of the Co-authorship of Countries and Authors’ Collaboration Involved in Sickle Cell Disease Research in Africa

The network analysis of co-authorship of countries not only provides an understanding of the social structure of the authors but also of the countries to which they belong. Figure 22 is the overlay visualization of co-authorship of countries involved in SCD research in Africa. It provides a visual representation of the research landscape for SCD in Africa. It also highlights the collaborative relationship between authors in countries involved in SCD research in Africa. The size of each node represents the number of publications authors from a country have co-authored with other authors. The color of each circle represents the year of co-authorship. A total of 112 countries were identified and out of which, 51 meet the threshold of minimum number of documents of 5. For each of the 51 countries, the total strength of the co-authorship links with other countries was calculated and the countries with the greatest total link strength were selected. From this, 50 countries scaled through and were used to construct the network map. A total of 7 distinct clusters were identified. Cluster 1 (red color) consisted of 15 countries including Algeria, Belgium, Cameroon, Democratic Republic of Congo. Cameroon is the country with the highest link strength of co-authorship in this cluster. Cluster 2 (green color) is made up of 13 countries. Some of them are; Kenya, South Africa, Uganda, Tanzania with the United States of America having the strongest link of co-authorship. Cluster 3 (blue color) had 12 countries. Among them, the United Kingdom had the highest link strength of co-authorship. Cluster 4 (yellow color) comprises 4 countries; Angola, Congo, Japan and Portugal. Cluster 5 (purple color) consisted of 4 countries; Brazil, Gambia, Kuwait and Nigeria. Cluster 6 (light blue) had Denmark and cluster 7 (orange) had Jamaica as the only country.

**Figure 22.**
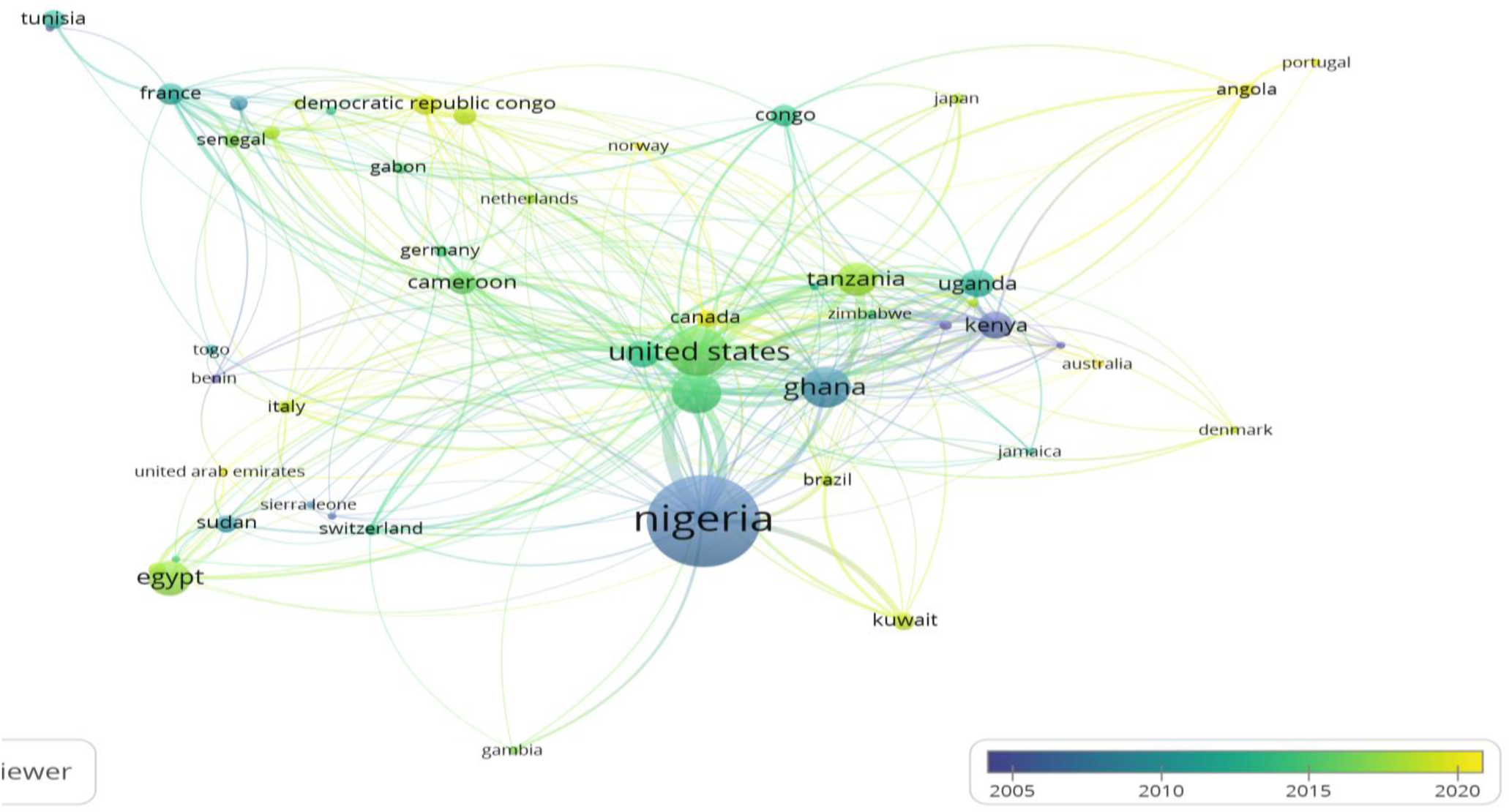
Overlay Visualization of Co-authorship by Country in Sickle Cell Disease Research in Africa.

Figure 23 shows how researchers in Africa collaborate on SCD research. Each circle represents an individual researcher and the line connecting each circle indicates that the two researchers have collaborated to co-author a publication on SCD in Africa. The size of a node represents the degree of collaboration associated with a researcher and each color cluster represents the research network that collaborates frequently. The collaboration between the various authors of publications on SCD in Africa is divided into 5 color clusters; blue, green, purple, red and orange. Authors with high degree of collaboration are located in the green and red color clusters respectively.

**Figure 23.**
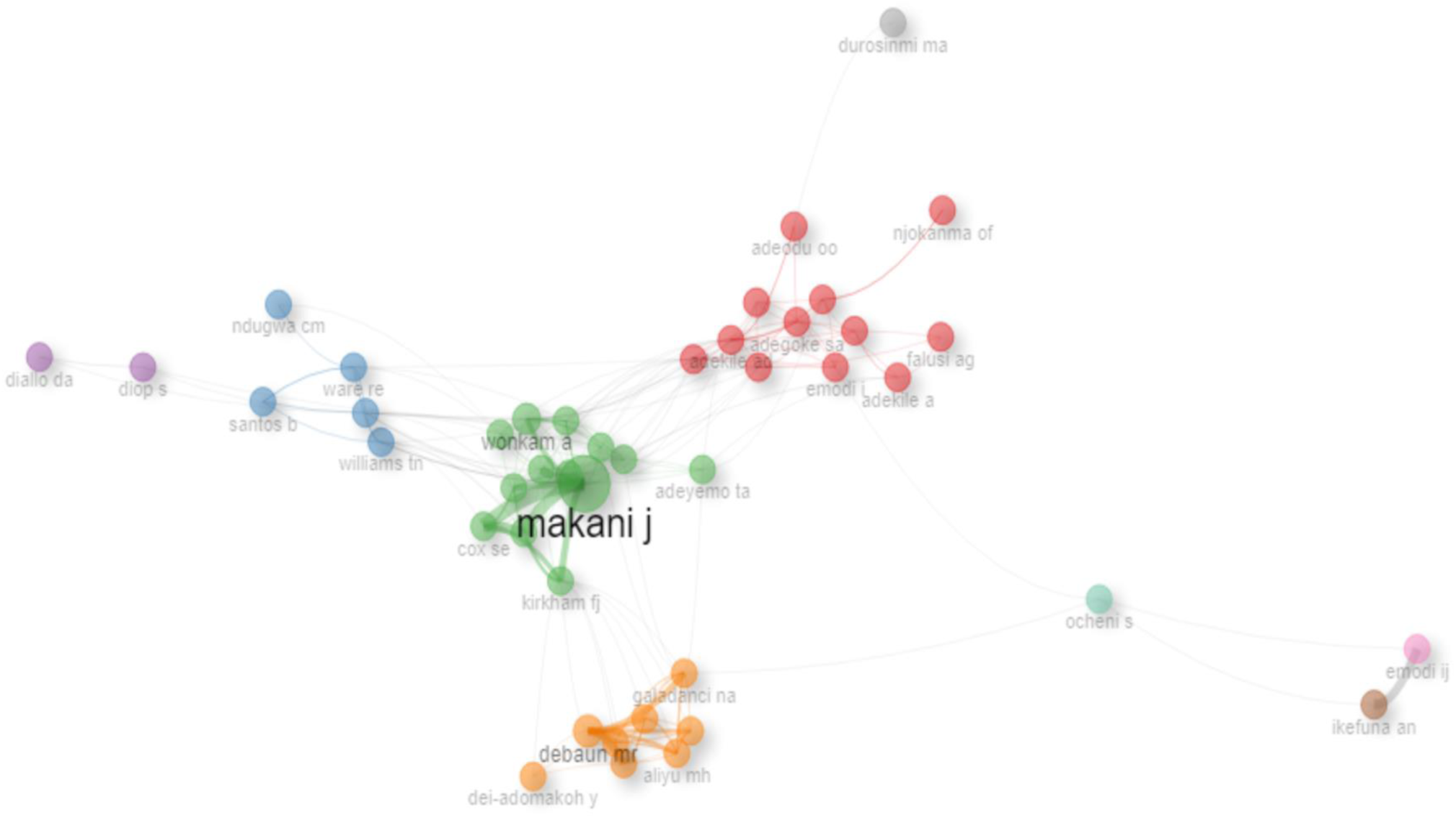
Author’s Collaboration Network in Sickle Cell Disease Research in Africa.

## DISCUSSION

Sickle cell disease is a life-threatening hematological disorder that has become a public health concern, especially in Africa. It is associated with a myriad of complications that affect every organ and system in the body. Africa bears the heaviest global burden of sickle cell disease. However, the visibility and impact of African-led research on the disease is not clearly defined. Therefore, conducting a systematic review and bibliometric visualization of publications on SCD in the context of Africa, will identify existing research outputs and trends. It will also provide evidence of the global contribution to research on SCD in Africa. The current study explored the research output on SCD in Africa using systematic review and bibliometric visualization approaches to provide a thorough overview of the research landscape of SCD in Africa.

Research on ‘sickle cell disease in Africa’ consisted of a total of 2132 publications contributed by 7178 authors across 728 journals. The most utilized document type in this study is the original research article which accounted for about 89.99% of all the document types. This is similar to the report of Musa *et al.,* (2021),and Borgohain *et al.,* (2022) which also reported that research articles are the most abundant document type. This is a reflection of the fact that the subject matter refers to clinical research and as such, the authors are more inclined to publish their research findings in research journals rather than other forms as these count for more visibility. Furthermore, original research articles serve as the primary means of disseminating new findings. On average, the annual growth rate of published literature on SCD in Africa is 4.77%. This annual growth rate shows that research on sickle cell disease in Africa is still lagging behind compared to a study conducted by Okoroiwu *et al*., (2020), which analyzed global SCD research from 1997 to 2017 and revealed that the growth of scientific publication output on SCD doubles every 4.52 years. Also, this growth is in the exponential stage with an annual increase of 4.93%.

In 1926, the first article on SCD in Africa was published by Archibald, R.G which he titled ‘A case of sickle cell anemia in Sudan’. Between 1926 to 1980, very few research output on SCD in Africa was recorded. This may be due to colonial rule as many of the African countries were still under colonial masters, as such, research was tailored to align with their interests. Also, during this period, much of the early research from Africa may have been published in non-English journals or local journals that were not indexed in international databases. Towards the end of the 20^th^ century and into the 21^st^ century, a surge in the number of published articles on SCD in Africa was observed. This may be due to the recent advances in technology, funding agencies and increased collaboration between African and global researchers.

The highest frequency of collaboration in research on SCD in Africa was observed to be between Niger Republic and Nigeria. This high collaboration frequency may be due to their close proximity as these two countries, located in West Africa, share a long border where there is free movement of people including researchers between them. Also, both Niger and Nigeria are significantly affected by the disease burden, thus making SCD research a shared priority. Furthermore, the highest MCP ratio was observed in Nigeria. This implies that researchers from Nigeria collaborate more with researchers from other countries. Also, this study revealed that Nigeria has published more research articles on SCD than any other Country in Africa. This is in tandem with the study of Musa *et al*., (2021) on global scientific research output on sickle cell disease which noted that Nigeria is the only African country that has published more than 300 articles on SCD.

In the last decade, bibliometric citation analysis has become popular in many fields of research as it estimates the influence of publications, authors, affiliations and journals through citation rates. Citations are often regarded as a measure of quality, with the assumption that if an article reports something important, then other researchers will make reference to it. An author or publication or institution is considered popular if it is heavily cited (Yari *et al*., 2020). Makani J of Mubimbili University of Health and Allied Sciences in Tanzania is the most cited author. This is an indication that this researcher is very popular when it comes to SCD in Africa. Also, based on total citation count, Nigeria is the most cited country. This is an indication of a strong research presence of Nigeria on SCD in Africa.

The h*-*index is a metric that attempts to measure both the productivity as well as the broad impact of an author’s cumulative research contribution (Rojas-Sánchez *et al.,* 2023). It takes into account both the number of articles published and the number of citations those publications have (Mondal *et al*., 2023). It gives a reliable assessment of the importance, significance and comprehensive impact of a researcher’s collective contribution to the body of knowledge (Agarwal *et al.,* 2015). It is calculated as the number of published articles by a researcher that have been cited at least ‘n’ times by other researchers (Hirsch, 2005). A high h*-*index suggests a significant impact. From this study, Makani J. had the highest impact with an h-index of 23. This implies that this author has 23 publications with at least 23 citations. Comparing authors based on their h-index alone may be misleading as it includes self-citations which can inflate the researcher’s impact and this may not accurately reflect their impact or influence in the field. Also, this matrix has often been criticized for favoring researchers who have been in the field for a longer period of time as they have been publishing for long, thus, have accumulated citations (Mondal *et al*., 2023). So, to compare the influence of authors in a particular field at different times, the m-index is considered. This index was designed to normalize the h-index, allowing for fair comparison between early-stage and late-stage researchers. The m-index for an author is calculated by dividing the individual author’s h-index by the number of years the author has been active in the field (Xu *et al.,* 2022). Hirsch suggested that an author with an m-index of 1 should be considered as a successful author while those with 2 or 3 should respectively be considered as outstanding or truly unique individuals. In this study, Makani. J, Williams., T.N., Cox S.E., Soka, D., Debaun, M.R, and Ware, R.E had an m-index of 1. Thus, they are considered as successful researchers on SCD in Africa. Debaun’s m-index was the highest which was 1.4 and has been publishing papers on SCD in Africa since 2015.

Thematic map divides research themes into two factors namely centrality and density (Nasir *et al*., 2020). The relevance degree (centrality) represented on the horizontal axis, indicates how relevant or central a theme is to the study. The vertical axis (development degree) suggests how much research has been conducted on a particular theme (Dastani, 2024). Based on this, the thematic map is divided into four quadrants. Themes at the upper right quadrant constitute the motor themes which represents the current focus of research on SCD in Africa. Although these themes are highly developed with high density, they however have low centrality (Katakidis *et al*., 2023). The presence of the keyword ‘sickle cell disease (scd)’ in this region could mean that this theme is currently the focus of research on SCD in Africa. Themes that appear at the lower right quadrant are the basic themes which represent the core concepts and foundational knowledge of SCD in Africa. Sickle cell disease, sickle cell anemia and antisickling are the themes on this quadrant. Thus, implying that they form the bedrock of research on SCD in Africa. At the upper left quadrant, is the niche theme. Although these themes are highly developed, they are however no longer explored. Keywords such as sickle cell trait, thalassemia, and priapism are located in this quadrant, thus, suggesting that research on them is of limited interest and no longer need to be investigated. The bottom left quadrant represents the emerging or declining themes. They are themes that can emerge (area of growing research interest) to be better or drop from research on SCD in Africa. Fetal hemoglobin, and oxidative stress are found in this quadrant. Keywords such as ‘anti-sickling’ are closely associated with ‘fetal hemoglobin’. This indicates that research on fetal hemoglobin reactivation is a promising avenue for developing anti-sickling therapies, as such, more research should be focused on it. Also, identification of biomarkers for SCD severity, genetic variation, gene therapy, hematopoietic stem cell transplantation and single cell analysis are some of the areas that have not been well explored in Africa.

Thematic evolution map shows how research themes on SCD in Africa have changed over a period of time. It shows the historical development of literatures on SCD in Africa (Nasir *et al*., 2020). Research on SCD in Africa was primarily focused on the disease itself and its associated complications. The period between 1926-2010 was dominated by topics such as ‘sickle cell disease’, ‘sickle cell anemia’ and ‘anti-sickling’. This suggests that a significant proportion of research on SCD during this period was focused on understanding the disease and exploring potential treatment options that can prevent the formation of hemoglobin polymers. The period between 2011-2019 saw a surge in a broader range of terms such as ‘epidemiology’, ‘inflammation’, ‘anti-sickling’ ‘quality of life’, ‘oxidative stress’ and ‘vasculopathy’. This signifies a growing interest in understanding the impact of the disease, complications, epidemiology, risk factors and underlying pathophysiological mechanism. As researchers continue to gain understanding of the pathophysiology of the disease, their focus shifted towards investigating specific complications and exploring treatment strategies. Between 2020-2024, research interests on SCD in Africa were focused on terms such as ‘anti-sickling’, ‘chronic kidney disease’, ‘blood transfusion’, and ‘fetal hemoglobin’. These suggest a growing interest in managing the complications associated with the disease. The recurring presence of ‘anti-sickling’ and ‘epidemiology’ across the periods is an indication that these areas are research hotspots in Africa.

A keyword is a significant part of a scholarly publication which plays an important role in research and information retrieval (Deka & Sarmah, 2020). It helps to improve the visibility of a research work. The keyword co-occurrence analysis ranked the most frequently occurring keywords as a means of identifying commonly studied topics and how these topics relate to each other (Belu & Marinoiu, 2025). During the analysis, a threshold of 5 keywords was chosen as the minimum number of times a keyword must appear. This implies that each of the total number of keywords identified for each unit of analysis of co-occurrence had appeared in at least 5 publications. Although there is no standard threshold for use in keyword co-occurrence analysis (Deka & Sarmah, 2020), however, the selection of a lower threshold will yield more keywords. The total link strength and occurrence where the matrix used to evaluate strength and frequency of occurrence of a keyword. The total link strength combines the frequency of occurrence with the number of other keywords with which a keyword occurs in the documents examined (Narong & Hallinger, 2023), while occurrence is the number of matches of keyword pairs obtained by direct search (Chigarev, 2024). The result of this analysis identified ‘Sickle cell anemia’ and ‘Sickle cell disease’ as the most frequently used keywords in SCD research in Africa. This is an indication that they are the primary focus of research on SCD in Africa. This is similar to the study of Musa *et al., (*2021) where ‘Sickle Cell Disease’ and ‘Sickle Cell Anemia’ are the most frequently used terms in global SCD research. SCD is an umbrella term that encompasses all forms of sickle hemoglobin disorders. SCA is the homozygous form of SCD which is the most common and severe form of the disease. These terms form the focus of research on SCD and as such, researchers tend to use them more in order to increase the visibility of their work.

The co-authorship analysis is a means of evaluating the relationships among authors, countries and organizations through the number of co-authored documents (Borgohain *et al.,* 2022). This study evaluated the co-authorship by countries from which Nigeria has co-authored more documents on SCD in Africa. This may be attributed to the high prevalence of the disease in this country which may be the drive for increased research activities.

The authors’ collaboration network revealed 5 color clusters which is an indication of the existence of various research groups. Researchers on the green color cluster are the most influential and collaborate with a large number of other researchers. Makani J. is the most influential researcher and has collaborated more with other researchers while Durosinmi, M.A., Diallo, D.A. and Njokanma, O.F. have fewer collaborations. This is an indication that they work more independently on SCD research in Africa. All researchers working on SCD in Africa must collaborate with each other and with those in the other parts of the world in order to better understand the disease in African context and also proffer ways in which to mitigate the disease burden and improve the quality of life of individuals with the disease.

## CONCLUSION

This study provided valuable information on the publication output and trend on SCD research in Africa from 1926 to 2024. Despite the disease prevalence in Africa, research on SCD in Africa is still lagging behind. A total of 2132 published articles on SCD in Africa were identified and these articles were authored by 7178 contributors across 718 journals with research articles as the most abundant document type. The highest number of published articles on SCD in Africa was recorded in the year 2022. The frequency of collaboration on SCD research in Africa was observed to be between Niger Republic and Nigeria. Nigeria is the most cited country and also has more corresponding authors of publications on research in Africa. Mubimbili University of Health and Allied Sciences in Tanzania is the most productive institution. Although, most of the productive institutions of research on SCD in Africa are located in Nigeria. Makani, J of Mubimbili University of Health and Allied Sciences in Tanzania is the most prolific and impactful researcher on SCD in Africa. Most of the works on SCD in Africa were published in East African Medical Journal whereas America Journal of Hematology is the most impactful source of published articles on SCD in Africa. Recent research on SCD in Africa is focused on investigating specific complications, disease management, and exploring various treatment strategies. Fetal hemoglobin, sickle cell disease and sickle cell anemia are the trending terms in research on SCD in Africa. The areas of growing interest in SCD research in Africa include fetal hemoglobin, inflammation and oxidative stress. The most used keywords in SCD research in Africa are Sickle cell disease and sickle cell anemia. To have a better understanding of the disease pathophysiology, and how the disease burden can be mitigated, African researchers working on SCD should collaborate more among themselves.

### Limitations and Future Research Direction

Despite the significance of this study, some limitations have been noted; the dataset used in this study was exclusively retrieved from the Scopus database while data from other relevant databases such as ProQuest, WoS, IEE were left out. The Scopus database alone cannot capture all relevant documents on the current discourse. Future studies can seek to integrate two or more of these databases. The data used in this study were exclusively sourced from English language works in the Scopus database and this may lead to the exclusion of other relevant works from other languages. Furthermore, the keyword strings used to retrieve the dataset may not be exhaustive. This may be due to the inherent challenge of identifying all relevant documents. As such, the need to develop a more comprehensive set of keywords strings. In future, the scope of this study can be widened by considering hemoglobinopathies as a whole and can also be refined to either focus on just a particular country or region within Africa.

## Data Availability

All data produced in the present study are available upon reasonable request to the authors

